# The time course, extent, and durability of improvement in depressive symptoms, daily function, and quality of life with three years of vagus nerve stimulation in markedly treatment-resistant major depression: A RECOVER study report

**DOI:** 10.64898/2026.07.31.26358854

**Authors:** Charles R. Conway, Scott T. Aaronson, A. John Rush, Ying-Chieh (Lisa) Lee, Charles Gordon, Olivia Shy, Mark T. Bunker, Patricio Riva-Posse, Mark S. George, John Zajecka, Ziad Nahas, David L. Dunner, João L. de Quevedo, Martijn Figee, Brian J. Mickey, Rebecca M. Allen, Kevin Reeves, Donald Bohnenkamp, Christopher L. Kriedt, Vasilis C. Hristidis, Charles F. Zorumski, Matthew Macaluso, Walter Duffy, Yvette Sheline, Gustavo Alva, Cristina Cusin, Jeffrey I. Bennett, Quyen Tran, Roger S. McIntyre, Richard Hamish McAllister-Williams, Harold A. Sackeim

## Abstract

**Background:** In markedly treatment-resistant depression, the RECOVER trial compared the time course, extent, and durability of benefits in groups randomized to three years of active VNS (Early-Active) or one year of sham VNS followed by two years of active VNS (Delayed-Active).

**Methods:** For 436 participants completing month-12 (M12) assessments (Early-Active N=221; Delayed-Active N=215), longitudinal mixed-effects models characterized trajectories of benefit over 36 months in categorical outcomes pooled across blinded depressive symptom and global improvement measures. Additional analyses examined the proportions benefiting and the degree and durability of benefits.

**Results:** The Early-Active group had greater odds of improvement during the RCT phase (OR=1.66; 95% CI=1.17–2.35; *P*=0.005). This between-group difference increased until M12 and then progressively narrowed upon VNS activation in the Delayed-Active group, approaching parity by M36. Across both groups, emergence of new beneficiaries plateaued after 24 months of active VNS, but with participants progressively achieving higher degrees of benefit thereafter. Following activation, the Delayed-Active group had significant improvement in all six categorical outcome measures. Durability was strong across depressive symptoms, function, and quality-of-life outcomes. For participants receiving active VNS throughout, durability of response was 70% from M12 to M36.

**Conclusions:** Active VNS was associated with improved benefits for both treatment groups over years 2 and 3. New beneficiaries emerged during the first two years of active VNS with continued progression to higher levels of benefit in the third year. These clinical benefits were highly durable. ClinicalTrials.gov identifier: NCT03887715.

## Introduction

In major depressive disorder (MDD), greater treatment-resistance is associated with a lower likelihood of benefiting acutely from subsequent antidepressant treatments and poorer durability when benefits are obtained (1–4). The time course for symptomatic improvement varies from hours for esketamine (5) or racemic ketamine (6, 7) to months or years for implanted vagus nerve stimulation (VNS) (8, 9). When improvement occurs over time, the extent of benefit can change in two ways: (1) participants may experience a greater degree of benefit over time (e.g., initial partial response progresses to response or remission), and (2) the proportion of participants with meaningful improvement may increase over time.

We recently found that both the degree of benefit and proportion of participants who improved increased during the second year of active VNS in RECOVER participants initially randomized to active, adjunctive VNS (10). Benefits included improvements in depressive symptoms, daily function, and quality of life (QoL). These findings confirmed that, in this population, improvement with VNS occurs over a time course often extending beyond one year.

Durability of benefit is of particular concern because relapse and recurrence rates are substantial with treatment-resistant depressions (TRD) that improve acutely with pharmacotherapy (1, 11), electroconvulsive therapy (ECT) (12–14), or accelerated transcranial magnetic stimulation (TMS) (15) despite continuation and maintenance therapies.

In contrast, ∼80% of participants who improved after one year of VNS maintained that degree of benefit over the second year for seven outcomes spanning depressive symptoms, daily function, and QoL(10). These results replicated previous observations (9, 16) and, for depressive symptoms, durability was uniformly strong when examined separately for participants with partial response, response, and remission at one year.

RECOVER participants with MDD were randomized to one year of blinded active VNS (Early-Active) or sham VNS (Delayed-Active) followed by active stimulation. This report evaluates the outcomes through the third year of the trial. We compared trajectories of symptomatic improvement of both groups during the randomized controlled trial (RCT) (Year 1) and follow-up (Years 2-3) phases. Outcomes in the Delayed-Active group after the Year 1 RCT phase have not been previously reported. In this group, with crossover to active stimulation at the end of Year 1, we expected an increase in both degree of benefit and number of participants benefiting during Year 2 and strong durability of benefits during Year 3 (second year of active VNS) paralleling findings in the Early-Active group.

We addressed the following questions:

1. *Trajectories of symptomatic improvement during the RCT and follow-up phases*: What were the trajectories of symptomatic improvement over the 36-month observation period in degree of benefit and percentage of participants benefiting? Did the randomized groups differ in these trajectories when separately considering the RCT and follow-up phases?
2. *Time course of improvement*: For individual measures of symptom severity, global improvement, function, and QoL, how did the extent of benefit (proportion of participants with benefit and the degree of benefit) change during post-RCT follow-up?
3. *Durability of benefit:* For each group, were the benefits achieved at the end of the first and second study years durable as assessed by the percentage of participants who maintained benefit over the subsequent one to two years?

## Methods and Materials

### Study Design

RECOVER, a multicenter, randomized, triple-blind, sham-controlled trial, evaluated the safety and efficacy of implanted cervical VNS over a 12-month period in participants with markedly TRD (ClinicalTrials.gov identifier: NCT03887715) (17). During the RCT phase (Year 1), participants received either active (N=249) or sham (N=244) VNS, both adjunctive to TAU. Thereafter, all participants received active VNS for four years, without constraint on VNS stimulation parameters or concomitant treatments. The blind was maintained throughout the trial. The study was approved by local institutional review boards at each of 84 participating sites. All participants provided written informed consent. Procedures conformed to the ethical standards of the Helsinki Declaration.

### Patient Sample

Participants (≥18 years of age) had nonpsychotic MDD based on DSM-5 criteria (18) and were either chronically depressed (≥2 years current major depressive episode [MDE]) or experienced ≥4 MDEs. They obtained insufficient benefit from ≥4 verified adequate antidepressant treatments during the current MDE (4), with no upper limit on number of failed treatments.

Inclusion in this report required completion of the end-of-RCT assessment (month [M] 12), which provided outcome data across the RCT phase and a baseline for evaluating follow-up change (M12-M36). The sample initially randomized to active (N=249) or sham (N=244) VNS included 15 participants in each group who withdrew before the first post-baseline assessment at M3. An additional 27 participants (Early-Active=13; Delayed-Active=14) did not complete M12 assessment. Thus, the analytic sample included 436 participants (Early Active=221; Delayed-Active=215). By M36, participants received either three (Early-Active) or two (Delayed-Active) years of stimulation.

### Assessment Intervals

During the RCT phase, symptom measures were obtained monthly from M3 to M12; daily function and QoL were assessed quarterly. From M12 to M36, all outcomes were assessed every 6 months.

### Outcome Measures

This report describes every depressive symptom, psychosocial function, and QoL measure collected in RECOVER. All benefit thresholds were defined *a priori*. Symptom measures completed by blinded, off-site raters via telephone interview included the Montgomery-Åsberg Depression Rating Scale (MADRS) (19) and Quick Inventory of Depressive Symptomatology– Clinician (QIDS-C) (20). Participants completed the Quick Inventory of Depressive Symptomatology–Self-Report (QIDS-SR) (20). Each participant’s blinded-clinician completed the Clinical Global Impression–Improvement (CGI-I) scale (21). Categorical depressive symptom and CGI-I outcomes were defined as no benefit, partial response, response, and remission **(Table 1)**. Depending on the statistical analysis, these outcomes were defined using inclusive definitions, where participants with higher levels of benefit are included in subgroups achieving lower levels (e.g., response includes remission) or exclusive definitions, where subgroups only contain participants who met a specific threshold of benefit and not those at higher levels.

**Table 1.** Definitions of outcome categories for each categorical outcome measure.

| Scale | Partial response | Response | Remission |
| --- | --- | --- | --- |
| MADRS | ≥30% reduction from baseline | ≥50% reduction from baseline | Score ≤9 |
| QIDS-C | ≥30% reduction from baseline | ≥50% reduction from baseline | Score ≤5 |
| QIDS-SR | ≥30% reduction from baseline | ≥50% reduction from baseline | Score ≤5 |
| CGI-I | Score ≤3 | Score ≤2 | Score=1 |
|  | **Meaningful benefit** |  |  |
| WPAI item 6 | ≥2-point reduction from baseline | — | — |
| Mini-Q-LES-Q | ≥11.89% increase from baseline | — | — |

No meaningful benefit (NB) was defined as not achieving at least a partial response or meaningful benefit.
These definitions of benefit subgroups are inclusive, as participants with higher degrees of benefit are automatically included in the subgroups with a lower degree of categorical benefit (e.g., remission included in response subgroup). However, some analyses track changes over time in the proportions of participants achieving specific ranges of benefit, such as alluvial plots representing within-participant transitions, such as from response to remission. These analyses use exclusive definitions where each benefit subgroup is restricted to a specified range of improvement (e.g., response without remission). The methods used in specific analyses specify whether outcomes were defined inclusively or exclusively.
Abbreviations: CGI-I, Clinical Global Impression–Improvement; MADRS, Montgomery-Åsberg Depression Rating Scale; QIDS-C, Quick Inventory of Depressive Symptomatology–Clinician; QIDS-SR, Quick Inventory of Depressive Symptomatology–Self-Report.

The impact of depression on daily activities during the previous week was measured with Item 6 of the Work Productivity and Activity Impairment Questionnaire (WPAI) (22–24) using a scale ranging from 0 (no effect) to 10 (completely prevented activities). Only item 6 was examined because 75% of participants were unemployed at baseline; the remaining WPAI items assess work productivity. For WPAI item 6, meaningful benefit (MB) was distinguished from no benefit (NB) and defined as a reduction of ≥2 points from baseline (25).

QoL was assessed with the 14-item Quality of Life Enjoyment and Satisfaction Questionnaire (Q-LES-Q) that measures the degree of enjoyment and satisfaction over a range of functional domains and the Mini-Q-LES-Q, a validated 7-item subset of the Q-LES-Q designed to assess QoL in depression (26). For both measures, MB was defined as ≥11.89% increase in score from baseline (27). Since findings were highly concordant, only Mini-Q-LES-Q results are presented.

The WHO Disability Assessment Schedule (WHODAS) 2.0 (28) evaluates functioning across six domains (cognition, mobility, self-care, getting along, life activities, and participation). The EuroQol Five-Dimension Five-Level Questionnaire (EQ-5D-5L) (29, 30) assesses QoL in mobility, self-care, usual activities, pain/discomfort, and anxiety/depression. Benefit thresholds have not been validated for EQ-5D Visual Analogue Scale or WHODAS 2.0 scores and change in raw scores was examined.

Our previously developed tripartite composite metric assessing breadth of benefit by counting the number of domains (symptoms, function, and QoL) in which participants improved (25, 31) was beyond the scope of this report.

## Statistical Analysis

Analyses were conducted for 8 outcome measures over 36 months. Confidence intervals (CIs) and *P*-values were not adjusted for multiplicity. Statistical significance was assessed using a two-sided α level of 0.05. All analyses were performed with SAS 9.4 (SAS Institute, Cary, NC).

### Clinical Outcome Trajectories Over Three Years: Overall Longitudinal Analyses

To characterize outcome trajectories during the RCT phase and subsequent two-year follow-up, an overall integrated (across 4 scales) longitudinal analysis of the extent of benefit was conducted. This analysis used a cumulative logit mixed-effects model applied to ordinal (exclusive) outcome categories (0=no benefit [NB], 1=partial response, 2=response, 3=remission) derived from MADRS, QIDS-C, QIDS-SR, and CGI-I assessments across all visits through M36. These four scales comprised the totality of the blinded mood and clinical function inventories administered and all four scales have established (a priori) ordinal thresholds (**Table 1**). Applying an overall longitudinal model across scales and measurement occasions enhanced reliability of outcome assessment and addressed multiplicity concerns (32, 33). The primary model treated assessment month as a continuous variable. A secondary analysis treated assessment month as a categorical variable to characterize visit-specific changes over time. The models’ fixed effects included treatment group (Early-Active vs. Delayed-Active), month, study phase (RCT vs. follow-up), and all interactions among these variables to assess whether improvement trajectories differed between treatment groups and study phases. Additional covariates included age (<65 vs. ≥65), baseline MADRS severity (<34 vs. ≥34), clinical scale, and a baseline score standardized for each scale. Subject-specific random intercepts were included to account for repeated measurements. Model estimates and contrasts were expressed as odds ratios (ORs). For within-group contrasts, ORs greater than 1 indicated greater improvement in outcome category (e.g., response vs. partial response) later in time. For between-group contrasts, ORs greater than 1 favoured the Early-Active group. Significant interactions in the overall model were explored through follow-up analyses of individual scales within treatment groups and study phases.

As a sensitivity analysis, the primary longitudinal analysis was repeated after redefining benefit as response or remission, combining partial response and NB into a single category. This analysis evaluated whether the primary findings were maintained when only response and remission were treated as clinically meaningful.

The proportions of participants achieving partial response, response, and remission (inclusive) across the four clinical scales were calculated at each assessment to determine whether longitudinal improvements resulted from progression to higher benefit categories, increases in the proportion of participants benefiting, or both.

## Extent of Benefit During Years 2 and 3

For each treatment group and outcome measure, alluvial plots displayed participant-level transitions among exclusive outcome categories at M12, M24, and M36. These plots were based on (1) a completer sample including participants with data at M12, M24, and M36, with no imputation of missing data, and (2) a last observation carried forward (LOCF) sample that retained all participants.

Given time- and group-dependent effects observed in the overall longitudinal analysis, cumulative logit mixed-effects models were separately applied to the six scales with categorical outcomes using data from all assessments. For the Mini-Q-LES-Q and WPAI item 6, longitudinal analyses examined binary outcomes (NB=0; MB=1). Forest plots displayed ORs and 95% CIs for within-group changes in degree of benefit. Similar mixed-effects models specifying a normal distribution were fit to the total continuous scores for each measure, including the EQ-5D-5L and WHODAS.

For each scale, McNemar’s exact test assessed changes in the proportions of participants across inclusive outcome categories at M12, M24, and M36. Corresponding 95% CIs for paired differences were estimated using the Mantel–Haenszel method. Changes in the proportion classified as NB quantified emergence of benefit during follow-up.

### Durability of Benefit

For the MADRS, QIDS-C, QIDS-SR, and CGI-I, durability was defined inclusively as the proportions of participants who met partial response, response, or remission criteria at M12 or M24 and continued to meet or exceed the same thresholds at subsequent assessments. For the WPAI item 6 and Mini-Q-LES-Q, durability was defined as maintenance of MB. Durability was also examined separately for participants with an insufficient response to ECT, TMS, or esketamine during the current MDE.

## Results

### Demographics, Illness Course, and Treatment History

Baseline demographic, clinical, and treatment history characteristics of the two groups were highly similar (**Table S1**). Participants averaged 17.7 years in current episode, with 53.1% of their lifetimes in depression. They had not benefited sufficiently from 13.3 lifetime antidepressant trials; 67.0% received ECT and/or TMS in the current MDE. Participant retention rates beyond Year 1 were similar for the two groups (83%–87% of M12 completers provided outcome assessments at M24; 74%–79% at M36; **Table S2**).

### Outcome Trajectories Over Three Years: Overall Longitudinal Analysis

In the overall longitudinal analysis combining data from the MADRS, QIDS-C, QIDS-SR, and CGI-I, extent of benefit increased with time, with significant effects of Study Phase (RCT vs follow-up), Month, and the interaction between Study Phase and Month (all *P’s*<0.0001; **Table S3** and **Figure S1**). Improvement across the four scales continued during both the RCT phase and the subsequent two-year follow-up.

The treatment groups differed in extent of improvement during the RCT phase (**Table S4**). During this period, the odds of achieving a more favorable outcome category increased by approximately 4.7% per month for the Early-Active group relative to the Delayed-Active group (*P*=0.0001). Across M3-M12, the Early-Active group demonstrated greater cumulative odds of improvement than the Delayed-Active group (OR=1.66, 95% CI 1.17–2.35, *P*=0.005). Significant between-treatment differences favoring the Early-Active group were observed from M5 through M12, with ORs increasing from 1.48 (95% CI, 1.04–2.11; *P*=0.031) at M5 to 2.04 (95% CI, 1.42– 2.93; *P*<0.001) at M12. Following device activation in the Delayed-Active group after M12 assessments, the between-group treatment effect progressively attenuated. The estimated OR decreased to 1.39 at M18 and reached parity by M36 (OR=1.05; 95% CI, 0.71–1.54).

**Figure 1a** presents cumulative ORs for between-group differences over the 3-year study period with values above 1 indicating advantage for the Early-Active group. These values derive from the primary logit mixed-effects model where time was a continuous linear variable. **Figure 1b** presents the same analysis with visit as a categorical variable, allowing treatment effects to vary with assessment visit. **Figures 1a and 1b** display the same phenomena: efficacy differences favoring Early-Active VNS increased throughout the RCT phase, peaking at M12. This treatment difference steadily narrowed following VNS activation in the Delayed-Active group.

**Figure 1.**
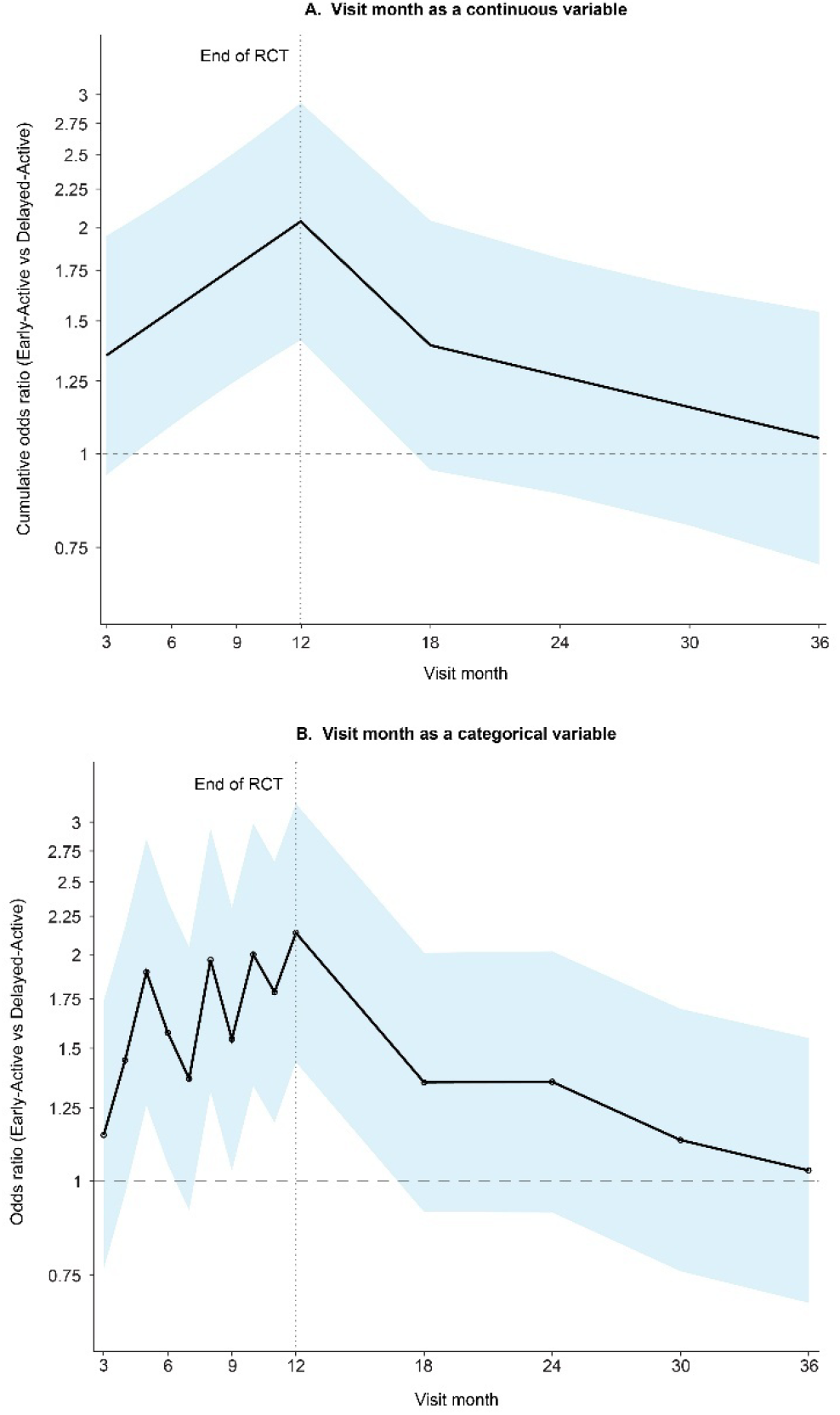
Cumulative odds ratios pooling data from the MADRS, QIDS-C, QIDS-SR, and CGI-I scales comparing Early-Active and Delayed-Active groups where **A.** visit was treated as a continuous variable, and **B.** visit was treated as a categorical variable. The first 12 months represent the RCT phase during which the Early-Active group received active VNS treatment and the Delayed-Active group received sham VNS. After Month 12, all participants received active VNS. Data are based on exclusive ordinal outcomes. The line represents the estimated odds ratio over time. The shaded area represents the range between the upper and lower confidence limits Abbreviations: RCT, randomized controlled trial.

The sensitivity analysis using only three mutually exclusive categories (NB, response, remission) produced similar findings, including significant advantage for Early-Active VNS during the RCT phase (OR=1.37; 95% CI, 1.03–1.81; *P*=0.03).

These overall longitudinal findings, indicating time-dependent, between-treatment group differences in extent of benefit, could reflect progression to higher benefit categories, increases in the proportion of participants benefiting, or both.

## Timing of Attainment of Outcome Thresholds: Early-Active vs Delayed-Active

**Figure 2** displays the mean proportion of participants meeting each level of benefit threshold averaged across MADRS, QIDS-C, QIDS-SR, and CGI-I at each visit. In the Early-Active group, the proportion of participants achieving partial response (**Figure 2A**) increased until M24 and plateaued over the remaining 12 months. In contrast, participants achieving response and remission (**Figures 2B**, **2C**) increased continuously over the entire 3-year period. Thus, most participants ultimately achieving benefit did so by M24, while more participants progressed to response and remission at later assessments.

**Figure 2.**
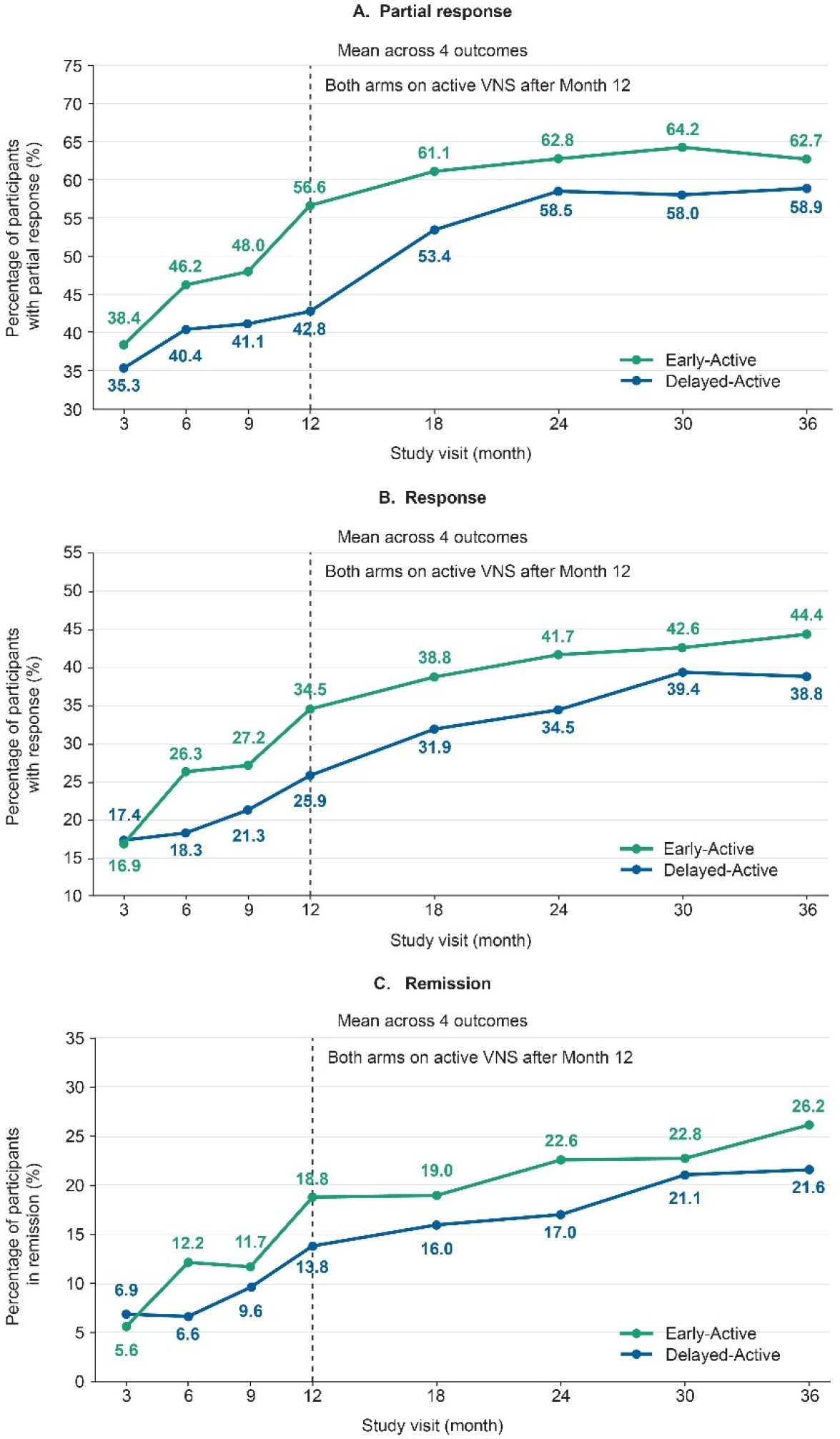
Mean proportion of participants meeting **A.** partial response, **B.** response, and **C.** remission across four clinical outcomes (MADRS, QIDS-C, QIDS-SR, CGI-I) from Months 3 to 36. Data presented are based on inclusive outcome definitions. Abbreviations: CGI-I, Clinical Global Impression–Improvement; MADRS, Montgomery-Åsberg Depression Rating Scale; QIDS-C, Quick Inventory of Depressive Symptomatology–Clinician; QIDS-SR, Quick Inventory of Depressive Symptomatology–Self-Report; VNS, vagus nerve stimulation.

The Delayed-Active group had a marked increase in the proportion with any benefit between M12 and M24 (**Figure 2A**) with few new beneficiaries thereafter. This increase, coincident with device activation, was greater than the concurrent increase in the Early-Active group. However, like the Early-Active group, participants achieving response and remission (**Figures 2B**, **2C**) progressively increased through the third year.

## Graphical representation of the emergence, loss, and maintenance of benefit

The alluvial diagrams (**Figure 3)** depict participant-level transitions among categorical outcomes on MADRS, QIDS-C, and CGI-I between M12 and M24, and M24 and M36 among participants with non-missing values at all three assessments. **Figure S2** presents alluvial plots for QIDS-SR, Mini-Q-LES-Q, and WPAI item 6. **Figures S3 and S4** present alluvial plots for the same 6 outcome measures using LOCF methods to examine potential effects of dropout.

**Figure 3.**
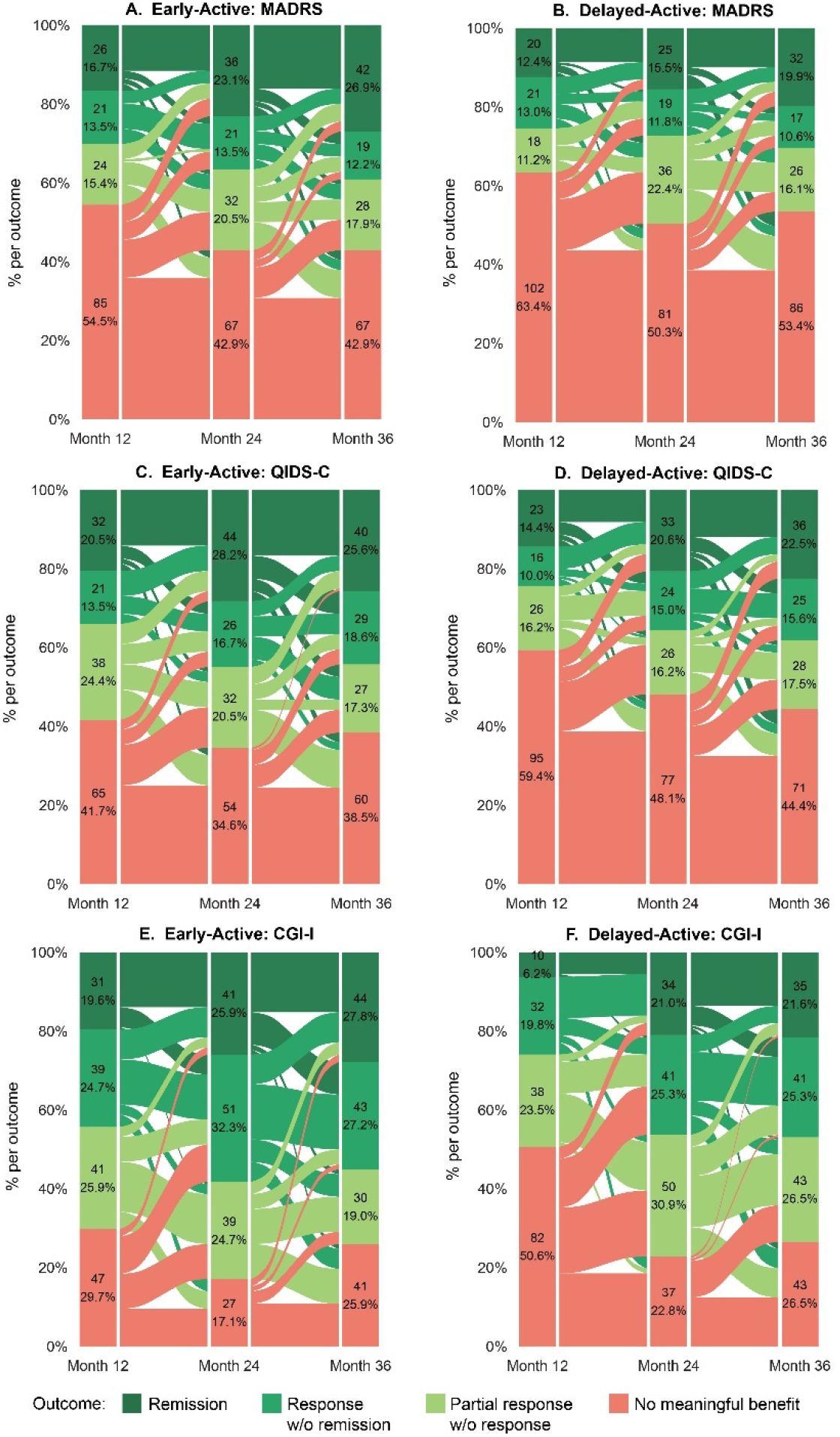
Alluvial diagrams of change in **A. B.** MADRS, **C. D.** QIDS-C, and **E. F.** CGI-I outcome categories in the Early-Active and Delayed-Active groups from Month 12 to Month 36. Data presented are based on exclusive outcome definitions. Data presented are samples with no missing values at the three time points. Abbreviations: CGI-I, Clinical Global Impression–Improvement; MADRS, Montgomery-Åsberg Depression Rating Scale; QIDS-C, Quick Inventory of Depressive Symptomatology–Clinician; QIDS-SR, Quick Inventory of Depressive Symptomatology–Self-Report.

Across measures, in both the Early-Active and Delayed-Active groups, the proportion of patients without benefit decreased between M12 and M24, as many participants maintained or improved benefit in the second year. Over the third year, the proportion achieving at least MB was largely unchanged, but many of those already benefiting transitioned to higher levels of benefit. Very few participants transitioned from a benefit category to NB during M24 or M36, indicating low rates of relapse.

Though similar, alluvial plots for Early-Active and Delayed-Active groups were not identical. The Delayed-Active group had greater emergence of benefit between M12 and M24 and, across the six categorical outcome measures, showed marked improvement during the 2-year follow-up.

Similar patterns characterized alluvial plots in the LOCF sample, suggesting that differential dropout was not a major determinant of these trajectories (**Figures S3 and S4**).

## Extent of Benefit During Years 2 and 3: Individual Measures

### Degree of Benefit in Years 2 and 3

**Table S5** presents raw data by scale and visit. Scale-specific mixed-effects models generally mirrored findings of the overall longitudinal analysis (**Tables S6 and S7**). Figure 4 summarizes within-group changes in degree of benefit from M12 to M24, and M12 to M36.

**Figure 4.**
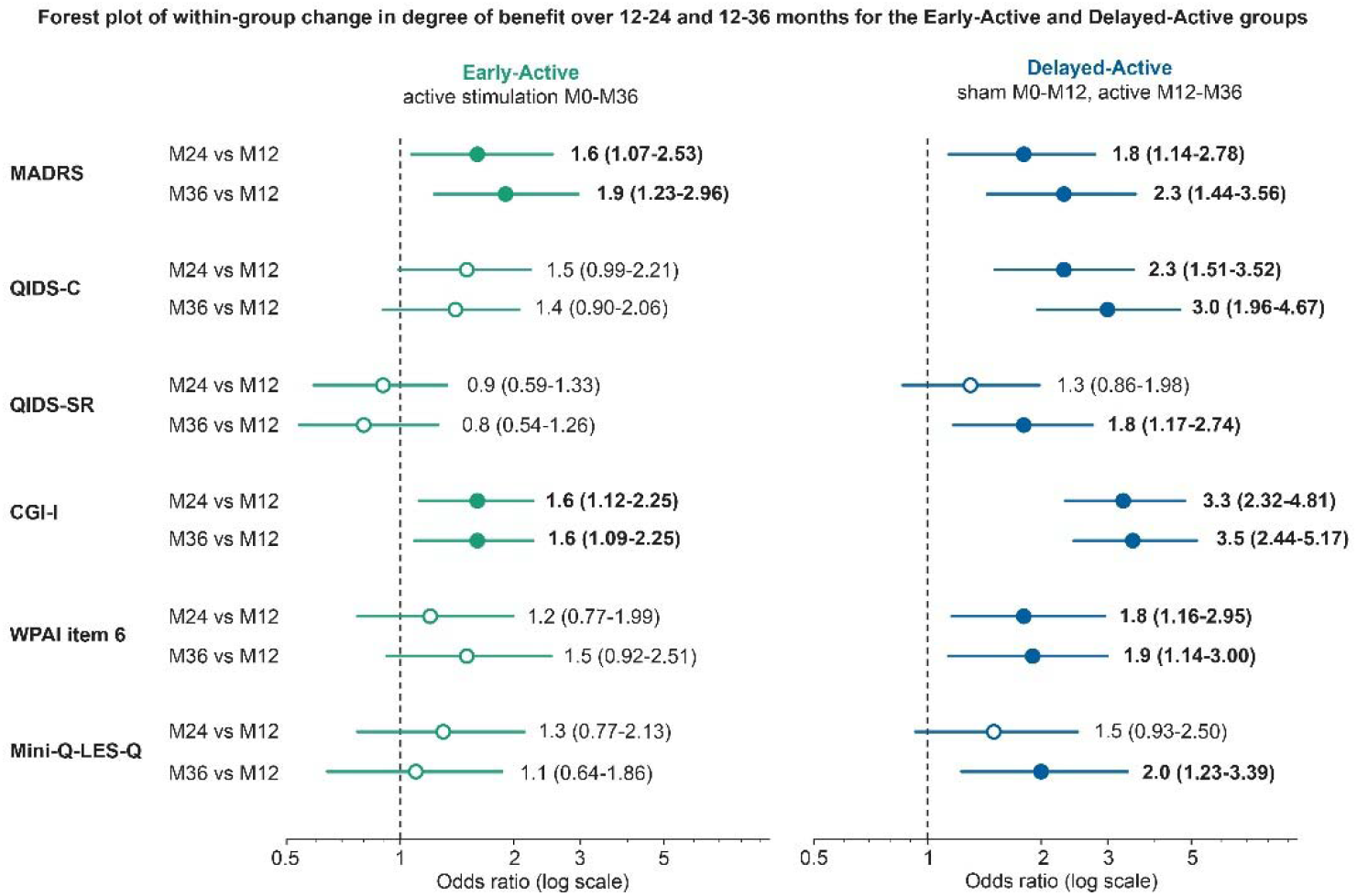
Forest plot of within-group change in extent of benefit over 12-24 and 12-36 months for the Early-Active and Delayed-Active groups. Data presented are based on exclusive ordinal outcome definitions. OR>1 indicates greater odds of improved status at the later visit. Contrasts are within-group over time and not comparisons between groups. Solid circles indicate statistically significant effect (p<0.05). WPAI item 6 and Mini-Q-LES-Q use a 2-level outcome status (0=no benefit, 1=MCID reached for meaningful benefit). All other measures use a 4-level outcome status (0=no benefit; 1=partial response without response; 2=response without remission; 3=remission). Abbreviations: CGI-I, Clinical Global Impression–Improvement; MADRS, Montgomery-Åsberg Depression Rating Scale; Mini-Q-LES-Q, 7-item subset of the Quality of Life Enjoyment and Satisfaction Questionnaire; QIDS-C, Quick Inventory of Depressive Symptomatology–Clinician; QIDS-SR, Quick Inventory of Depressive Symptomatology–Self-Report; VNS, vagus nerve stimulation; WPAI, Work Productivity and Activity Impairment Questionnaire.

Following device activation, the Delayed-Active group showed a significant increase in degree of benefit from M12 to M24 in 4 of 6 individual measures and over the full two-year follow-up period (M12 to M36) in all six measures (**Figure 4**). For example, with the CGI-I, the Delayed-Active group was more than 3 times more likely to have an improved outcome category (e.g. remission vs. response) at M24 relative to M12. In contrast, the Early-Active group showed significant improvement in the degree of benefit from M12 to M24 only for the MADRS and CGI-I. Neither group demonstrated significant changes on any individual measure from M24 to M36.

The pattern of significant group changes in raw scores mirrored effects on categorical outcomes (**Table S7**). From M12 to M36, the Delayed-Active group improved significantly on seven of eight raw-score measures, including the WHODAS, while the Early-Active group improved on the MADRS, QIDS-C, and CGI-I.

## Proportion of Participants Who Benefited during Years 2 and 3

**Table S8** summarizes the proportion of participants within each treatment group who achieved each categorical level of benefit on each measure during follow-up. Over the two-year follow-up (M12 to M36), the proportion of Delayed-Active participants achieving partial response increased significantly on all six measures. Significant gains in response and remission were also observed on selected depressive symptom and clinical impression measures. These improvements were driven primarily by increases from M12 to M24, as none of the within-group comparisons from M24 to M36 were statistically significant.

In the Early-Active group, significant increases in the proportions achieving partial response, response, and remission were observed on selected measures during follow-up. As in the Delayed-Active group, these gains occurred primarily from M12 to M24, with improvements that were not statistically significant during M24 to M36.

## Durability of Benefit

**Table 2** presents durability findings for the two groups for all measures at M24 and M36, organized by degree of benefit at M12 or M24. The median percentage of participants who achieved and maintained partial response or MB (or better) was 81.1% for the Early-Active group and 81.5% for the Delayed-Active group between M12 and M24, 78.6% and 78.2% between M24 and M36, and 76.7% and 75.5% between M12 and M36. Participants who attained response or remission maintained partial response at higher rates. Durability remained high even when maintenance of response or remission was required.

**Table 2.** Percentage of participants who maintained meaningful benefit or partial response, response, or remission at Months 24 and 36 for the **A.** Early-Active and **B.** Delayed-Active groups.^a^

**A. Early Active**
|  | Maintenance of at least partial response |  |  |
| --- | --- | --- | --- |
| Outcome Measure | In participants with at least partial response at Month 12 |  | In participants with at least partial response at Month 24 |
|  | At least partial response at Month 24 | At least partial response at Month 36 | At least partial response at Month 36 |
| MADRS | 82.9% (68/82) | 77.2% (61/79) | 78.7% (70/89) |
| QIDS-C | 80.8% (84/104) | 75.8% (75/99) | 78.4% (80/102) |
| QIDS-SR | 79.6% (90/113) | 69.5% (73/105) | 72.7% (72/99) |
| CGI-I | 85.3% (110/129) | 79.2% (95/120) | 81.7% (107/131) |
| Function (WPAI item 6)<sup>b</sup> | 79.2% (80/101) | 80.8% (80/99) | 79.4% (81/102) |
| QoL (Mini-Q-LES-Q)<sup>b</sup> | 81.3% (91/112) | 76.2% (77/101) | 76.9% (80/104) |
| Median | 81.1% | 76.7% | 78.6% |
|  | Maintenance of at least partial response |  |  |
| Outcome Measure | In participants with at least response at Month 12 |  | In participants with at least response at Month 24 |
|  | At least partial response at Month 24 | At least partial response at Month 36 | At least partial response at Month 36 |
| MADRS | 92.6% (50/54) | 88.0% (44/50) | 86.0% (49/57) |
| QIDS-C | 90.3% (56/62) | 89.8% (53/59) | 90.0% (63/70) |
| QIDS-SR | 87.3% (62/71) | 77.6% (52/67) | 81.7% (58/71) |
| CGI-I | 92.2% (71/77) | 84.2% (64/76) | 89.1% (82/92) |
| Median | 91.3% | 86.1% | 87.6% |
|  | Maintenance of at least response |  |  |
| Outcome Measure | In participants with at least response at Month 12 |  | In participants with at least response at Month 24 |
|  | At least response at Month 24 | At least response at Month 36 | At least response at Month 36 |
| MADRS | 74.1% (40/54) | 68.0% (34/50) | 71.9% (41/57) |
| QIDS-C | 79.0% (49/62) | 72.9% (43/59) | 70.0% (49/70) |
| QIDS-SR | 74.6% (53/71) | 62.7% (42/67) | 60.6% (43/71) |
| CGI-I | 83.1% (64/77) | 71.1% (54/76) | 77.2% (71/92) |
| Median | 76.8% | 69.6% | 71.0% |
|  | Maintenance of remission |  |  |
| Outcome Measure | In participants with remission at Month 12 |  | In participants with remission at Month 24 |

|  | **Remission at**<br>**Month 24** | **Remission at**<br>**Month 36** | **Remission at**<br>**Month 36** |
| --- | --- | --- | --- |
| MADRS | 65.5% (19/29) | 64.3% (18/28) | 69.4% (25/36) |
| QIDS-C | 61.1% (22/36) | 52.9% (18/34) | 59.1% (26/44) |
| QIDS-SR | 47.8% (22/46) | 46.7% (21/45) | 65.8% (25/38) |
| CGI-I | 69.7% (23/33) | 54.5% (18/33) | 58.5% (24/41) |
| Median | 63.3% | 53.7% | 62.5% |

**B. Delayed- Active**
|  | Maintenance of at least partial response |  |  |
| --- | --- | --- | --- |
| Outcome Measure | In participants with at least partial response at Month 12 |  | In participants with at least partial response at Month 24 |
|  | At least partial response at Month 24 | At least partial response at Month 36 | At least partial response at Month 36 |
| MADRS | 82.4% (56/68) | 70.0% (42/60) | 70.0% (56/80) |
| QIDS-C | 78.7% (59/75) | 71.2% (47/66) | 77.1% (64/83) |
| QIDS-SR | 81.1% (73/90) | 77.5% (62/80) | 79.3% (73/92) |
| CGI-I | 90.5% (76/84) | 79.3% (65/82) | 81.6% (102/125) |
| Function (WPAI item 6)<sup>b</sup> | 81.8% (72/88) | 73.4% (58/79) | 73.9% (68/92) |
| QoL (Mini-Q-LES-Q)<sup>b</sup> | 77.1% (64/83) | 78.1% (57/73) | 80.5% (66/82) |
| Median | 81.5% | 75.5% | 78.2% |
|  | Maintenance of at least partial response |  |  |
| Outcome Measure | In participants with at least response at Month 12 |  | In participants with at least response at Month 24 |
|  | At least partial response at Month 24 | At least partial response at Month 36 | At least partial response at Month 36 |
| MADRS | 85.7% (42/49) | 68.3% (28/41) | 77.3% (34/44) |
| QIDS-C | 84.4% (38/45) | 72.5% (29/40) | 82.5% (47/57) |
| QIDS-SR | 94.4% (51/54) | 89.6% (43/48) | 92.2% (47/51) |
| CGI-I | 88.4% (38/43) | 81.0% (34/42) | 85.3% (64/75) |
| Median | 87.1% | 76.8% | 83.9% |
|  | Maintenance of at least response |  |  |
| Outcome Measure | In participants with at least response at Month 12 |  | In participants with at least response at Month 24 |
|  | At least response at Month 24 | At least response at Month 36 | At least response at Month 36 |
| MADRS | 63.3% (31/49) | 56.1% (23/41) | 63.6% (28/44) |
| QIDS-C | 75.6% (34/45) | 65.0% (26/40) | 73.7% (42/57) |
| QIDS-SR | 68.5% (37/54) | 79.2% (38/48) | 70.6% (36/51) |
| CGI-I | 81.4% (35/43) | 69.0% (29/42) | 76.0% (57/75) |
| Median | 72.1% | 67.0% | 72.2% |
|  | Maintenance of remission |  |  |
| Outcome Measure | In participants with remission at Month 12 |  | In participants with remission at Month 24 |
|  | Remission at Month 24 | Remission at Month 36 | Remission at Month 36 |
| MADRS | 65.4% (17/26) | 65.0% (13/20) | 64.0% (16/25) |
| QIDS-C | 50.0% (14/28) | 58.3% (14/24) | 57.6% (19/33) |
| QIDS-SR | 58.1% (18/31) | 63.0% (17/27) | 68.0% (17/25) |
| CGI-I | 90.9% (10/11) | 70.0% (7/10) | 64.7% (22/34) |
| Median | 61.8% | 64.0% | 64.4% |

<sup>a</sup>Outcome definitions were inclusive.
<sup>b</sup>Partial response equates to clinically meaningful benefit for these outcome measures.
Abbreviations: CGI-I, Clinical Global Impression–Improvement; MADRS, Montgomery-Åsberg Depression Rating Scale; MB, meaningful benefit; Mini-Q-LES-Q, 7-item subset of the Quality of Life Enjoyment and Satisfaction Questionnaire; QIDS-C, Quick Inventory of Depressive Symptomatology–Clinician; QIDS-SR, Quick Inventory of Depressive Symptomatology–Self-Report; WPAI, Work Productivity and Activity Impairment Questionnaire.

Durability was consistently high across depressive symptom, clinical global impression, daily function, and QoL measures (**Figure 5)**. Durability tended to be greatest over shorter follow-up intervals (M12 to M24, and M24 to M36) but remained robust across the full two-year interval (M12 to M36). **Table S9** presents the comparison of durability rates for participants who did not benefit from interventional treatments (ECT, TMS, esketamine) in the current depressive episode compared to the total sample. Strong durability of benefit at 36 months was observed for both the Early-Active interventional nonresponders (medians: 62% interventional, 70% total) and Delayed-Active interventional nonresponders (medians: 64% interventional, 67% total).

**Figure 5.**
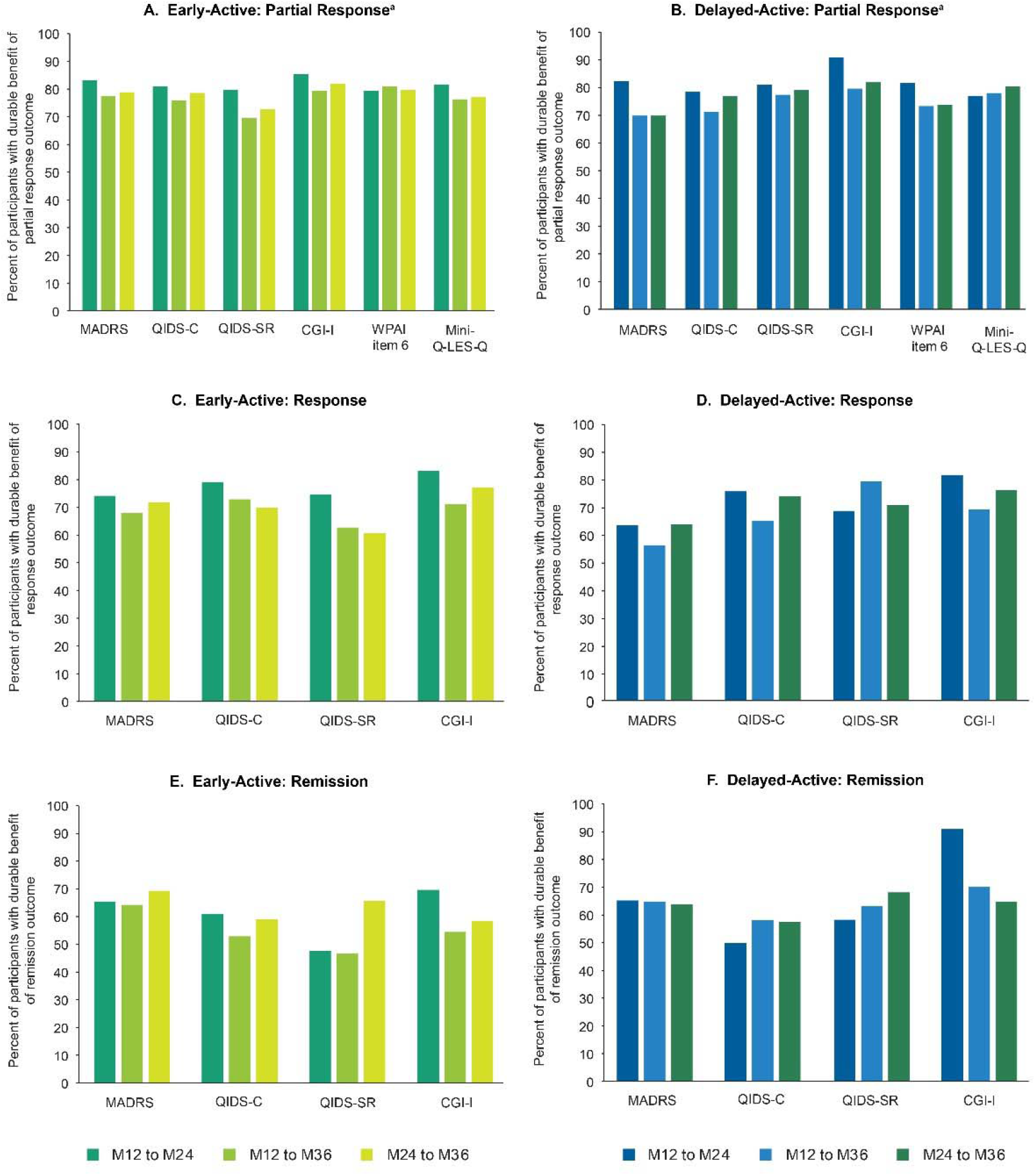
Durability of **A. B.** at least partial response, **C. D.** at least response, and **E. F.** remission across all measures in the Early-Active and Delayed-Active groups from Months 12 to 24, 12 to 36, and 24 to 36. Data presented are based on inclusive outcome definitions. ^a^For WPAI item 6 and Mini-Q-LES-Q, partial response equates to meaningful benefit. For WPAI item 6 and Mini-Q-LES-Q, partial response equates to meaningful benefit. Abbreviations: CGI-I, Clinical Global Impression–Improvement; MADRS, Montgomery-Åsberg Depression Rating Scale; MB, meaningful benefit; QIDS-C, Quick Inventory of Depressive Symptomatology–Clinician; QIDS-SR, Quick Inventory of Depressive Symptomatology–Self-Report.

## Discussion

RECOVER trial participants had markedly chronic and treatment-resistant depressions, features that predict a very low likelihood of attaining and sustaining meaningful clinical benefits with subsequent interventions. Over a 36-month period, we evaluated the time course, extent, and durability of clinical improvement in Early-Active and Delayed-Active randomized treatment groups. An overall longitudinal analysis of blinded categorical outcomes across multiple depressive symptom severity and global impression scales documented treatment-group differences in the trajectories of symptomatic improvement. The Early-Active group showed greater cumulative improvement during the RCT phase (months 3-12) compared to the

Delayed-Active group that received sham VNS during this phase. During the follow-up phase (months 12-36), these treatment-group differences reversed, as the Delayed-Active group demonstrated greater incremental improvement, and the magnitude of between-group difference in outcomes progressively attenuated.

These changes in extent of benefit could reflect differences over time in the *degree of benefit* achieved and/or *in the number of participants benefiting.* Both treatment groups had substantial increases in the percentage of participants with partial response until 24 months of active stimulation, with few new beneficiaries thereafter. In contrast, degree of benefit improved in both groups over the entire 36-month period as increasing percentages moved from partial response to response or remission. Consistent with previous studies (9, 34), these findings indicate that it may take one to two years to identify markedly TRD patients who benefit from VNS. Those who attain clinical benefit are likely to show further improvement over a 3-year treatment period.

The direction of treatment-group differences in degree of benefit and proportion of beneficiaries reversed at the end of the RCT phase, coincident with device activation in participants receiving sham VNS in Year 1. This pattern was anticipated, consistent with active VNS having greater impact on clinical outcomes than sham VNS. Furthermore, the time course and extent of benefit with active VNS was consistent across depressive symptoms, daily function and QoL outcomes.

Sustaining benefit once achieved is a fundamental challenge in markedly TRD, as high rates of relapse are observed when benefit results from pharmacological and other neuromodulatory interventions (1, 11–15). Across the two treatment groups, we found that 65–80% of those who benefited retained benefits over one and two years. For example, using a median across four measures (CGI-I, MADRS, QIDS-C, QIDS-SR) and response as the threshold, 77% of the Early-Active group retained response status between M12-24 and 70% between M12-36. In the Delayed-Active group these values were 72% for M12-24 and 67% for M12-36.

Prior studies document similar durability findings. In the continuation phases of the initial pilot trial (35) and original pivotal trial of VNS in TRD (34, 36), Sackeim et al. (16) found response at 12 months was maintained for 76% and 81% at 24 months for the pilot (30) and pivotal (31) trials, respectively. Kumar et al. (37), using a large VNS registry, found that patients receiving VNS plus TAU had longer times to relapse than patients receiving only TAU. Furthermore, in the current study, a sub-analysis documented high durability rates in participants with nonresponse to interventional treatments (ECT, TMS, esketamine) in the current episode. Thus, high likelihood of maintaining benefit over at least one to two years characterizes VNS in individuals with markedly TRD.

Clinical improvement with VNS occurs over a longer time than other treatments. At least two years of active VNS may be required to determine which individuals obtain at least partial benefit. A third year may be required to observe maximal degree of benefit. These time courses for onset and peak benefit are similar to VNS in medication-resistant epilepsy, where most beneficiaries are identified within two years, but a minority acquires benefit after three or four years (38).

## Limitations

Requiring 12-month completion to define the analytic sample potentially introduces concerns about selection bias. However, sample loss in year 1 was minimal (39), with high retention rates thereafter, and no group differences in data loss (**Table S2**: 85% of participants with 12-month data provided 2-year data; 76% provided 3-year data). After M12, changes in concomitant treatments were unconstrained and not examined here. However, our previous report on two years outcomes in the Early-Active group found no association between changes in concomitant treatments and benefits (10). The reversal of improvement trajectories after M12 was compatible with, but not determinative of, an active effect of VNS, as timing of the crossover was conveyed in the study consent.

## Conclusions

Clinical improvement with VNS unfolds over months to years, as more participants benefit and the average degree of benefit increases. With activation of VNS after one year of sham stimulation, there was a significant increase in the extent of benefit—mirroring the pattern of benefit observed with the Early-Active group during the RCT phase. The benefits achieved were highly durable over one and two years and pertained to symptoms, daily function, and QoL.

## Supporting information

List of IRBs

## Supplementary Material

Separate file.

## Acknowledgments

This work was supported by LivaNova, PLC, the developer and manufacturer of the Vagus Nerve Stimulation therapy system. Conducting the study, analyzing the data, and drafting the report were supported by LivaNova, PLC. Conducting the study was also supported by the Centers for Medicare & Medicaid Services. Final approval of the content of this manuscript and the decision to submit it were determined solely by the authors.

The authors are deeply grateful to the patients and their families for participating in the RECOVER study. We greatly appreciate the Centers for Medicare & Medicaid Services for providing financial support for the Vagus Nerve Stimulation therapy system devices and implantation surgeries. The authors would also like to thank Zeeba Kabir, PhD, and Paul Cao, PhD, of Simpson Healthcare, for their editorial assistance in accordance with the Good Publication Practice 2022 guidelines. This support was funded by LivaNova.

We would also like to thank the members of the RECOVER study group: Advanced Mental Health Care, Palm Beach, FL (Aron Tendler); Alivation Research, Lincoln, NE (Walter Duffy); AMR Baber Research, Naperville, IL (Riaz Baber); APG Research, Orlando, FL (Morteza Nadjafi); ATP Clinical Research, Costa Mesa, CA (Gustavo Alva); Barnes-Jewish Hospital, St Louis, MO (Donald Bohnenkamp); Beacon Medical Group Behavioral Health South Bend, South Bend, IN (Suhayl Nasr); Carilion Clinic, Roanoke, VA (Anita Kablinger); Center for Anxiety and Depression, Mercer Island, WA (David Dunner); Center for Neuropsychiatry and Brain Stimulation, Durham, NC (Sandeep Vaishnavi); Charak Center for Health and Wellness, Garfield Heights, OH (Rakesh Ranjan); DENT Neurologic Institute, Amherst, NY (Horacio Capote); Emory University, Atlanta, GA (Patricio Riva Posse IV); Florida Behavioral Medicine, Largo, FL (Ashok Patel); Florida Center for TMS, Orlando, FL (Todd Broder); Florida Center for TMS, St Augustine, FL (Heather Luing); Galiz Research, Hialeah, FL (Jose Gamez); Hapworth Research, New York, NY (William Hapworth); Healthy Perspectives, Nashua, NH (Hisham Hafez); Icahn School of Medicine at Mount Sinai, New York, NY (James Murrough); Kaizen Brain Center, La Jolla, CA (Mohammed Ahmed); Marshall Psychiatry, Huntington, WV (Suzanne Holroyd); Massachusetts General Hospital, Boston, MA (Cristina Cusin); Medical College of Georgia at Augusta University, Augusta, GA (Peter Rosenquist); Medical University of South Carolina, Charleston, SC (Mark George); Michigan Clinical Research Institute, Ann Arbor, MI (Rajaprabhakaran Rajarethinam); Mindful Behavioral Health, Boca Raton, FL (Ivan Cichowicz); Neuropsychiatric Associates at Woodstock Research Center, Woodstock, VT (Susan Smiga); NeuroScience and TMS Treatment Center, Brentwood, TN (Jonathan Becker); Northwest Behavioral Research Center, Marietta, GA (Michael Banov); Nova Psychiatry, Orlando, FL (David Medina); Offices of Psychiatry & Counseling Services, Moosic, PA (Matthew Berger); Ohio State University, Columbus, OH (Kevin Reeves); OU Physicians, Tulsa, OK (Ondria Gleason); Precise Research Centers, Flowood, MS (Joseph Kwentus); PsychCare Consultants Research, St Louis, MO (Mohd Malik); Psychiatry Care and Research Center, O’Fallon, MO (John Canale); Rush University Medical Center, Chicago, IL (John Zajecka); Seattle Neuropsychiatric Treatment Center, Seattle, WA (Rebecca Allen); SF-Care, San Rafael, CA (Jason Bermak); Sheppard Pratt Health Systems, Baltimore, MD (Scott Aaronson); Signature Research Associates, Fairlawn, OH (Anand Chaturvedi); Southern Illinois University School of Medicine, Springfield, IL (Jeffrey I. Bennett); Stedman Clinical Trials, Tampa, FL (Mary Stedman); Stony Brook University Hospital, Stony Brook, NY (Lucian Manu); Syrentis Clinical Research, Santa Ana, CA (John Duffy); Texas Tech University Health Science Center, El Paso, TX (Peter Thompson); Trinity Medical, Lewiston, NY (Alfred Belen III); UC San Diego, San Diego, CA (Mounir Soliman); University of Alabama Heersink School of Medicine, Birmingham, AL (Matthew Macaluso); University of Alabama Huntsville Regional Medical Center, Huntsville, AL (Richard Shelton); University of Minnesota, Minneapolis, MN (Ziad Nahas); University of Missouri, Columbia, MO (Muaid Ithman); University of Utah Neuropsychiatric Institute, Salt Lake City, UT (Brian Mickey); University of Wisconsin, Madison, WI (Steven Garlow); UPenn Perelman School of Medicine, Philadelphia, PA (Yvette Sheline); USC Keck School of Medicine, Los Angeles, CA (Ashraf Elmashat); UT Dell Medical School, Austin, TX (Julie Farrington); and UT McGovern Medical School, Houston, TX (João Quevedo).

## Previously published

The revised manuscript represents a substantial reworking of the version submitted to the preprint server (Conway CR, et al. medRxiv 2026; https://doi.org/10.64898/2026.07.31.26358854) shifting the manuscript from a descriptive long-term follow-up report to a more rigorous analysis of the time course, extent, and durability of benefit with vagus nerve stimulation (VNS) in treatment-resistant depression. The abstract, introduction, methods, results, and discussion have been extensively rewritten to emphasize when benefits emerge, how they progress over time, and how group differences evolve after activation of VNS. A major conceptual change is the introduction of pooled longitudinal mixed-effects analyses comparing Early-Active and Delayed-Active participants across the entire study period, allowing direct characterization of treatment trajectories and between-group differences over time. The revised manuscript highlights a key new finding that the Early-Active group demonstrated significantly greater improvement compared with the Delayed-Active group during the randomized phase, with differences peaking at Month 12 and narrowing after VNS was activated in the Delayed-Active group.

Methodologically, the revised manuscript removes the tripartite composite endpoint that was included in the preprint server version and instead focuses only on measures of depressive symptoms, global improvement, function, and quality of life. New overall longitudinal trajectory analyses, sensitivity analyses, updated figures, and revised durability assessments were added, while a number of detailed within-group, exploratory, and tripartite composite-endpoint discussions were condensed or removed. The discussion was also substantially refocused, placing greater emphasis on the clinical implications of delayed response to VNS, increasing depth of benefit over time, and the durability of VNS effects. Collectively, the revisions create a more cohesive scientific story that active VNS produces benefits that emerge gradually, deepen over several years, and remain highly durable in a markedly treatment-resistant population, while providing stronger statistical support for the distinct trajectories observed in the Early-Active and Delayed-Active groups.

## Author Contribution Statement

**Conceptualization:** Charles R. Conway, Scott T. Aaronson, A. John Rush, Ying-Chieh (Lisa) Lee, Charles Gordon, Olivia Shy, Mark T. Bunker, Quyen Tran, and Harold A. Sackeim; **Data curation:** Ying-Chieh (Lisa) Lee, Charles Gordon, Olivia Shy, and Quyen Tran; **Formal analysis:** Charles R. Conway, Scott T. Aaronson, A. John Rush, Ying-Chieh (Lisa) Lee, Charles Gordon, Olivia Shy, Mark T. Bunker, and Harold A. Sackeim; **Funding acquisition:** Mark T. Bunker; **Investigation:** Charles R. Conway, Scott T. Aaronson, A. John Rush, Ying-Chieh (Lisa) Lee, Charles Gordon, Olivia Shy, Mark T. Bunker, Patricio Riva-Posse, Mark S. George, John Zajecka, Ziad Nahas, David L. Dunner, João L. de Quevedo, Martijn Figee, Brian J. Mickey, Rebecca M. Allen, Kevin Reeves, Donald Bohnenkamp, Christopher L. Kriedt, Vasilis C. Hristidis, Matthew Macaluso, Walter Duffy, Yvette Sheline, Gustavo Alva, Cristina Cusin, Jeffrey I. Bennett, Quyen Tran, and Harold A. Sackeim; **Methodology:** Charles R. Conway, Scott T. Aaronson, A. John Rush, Ying-Chieh (Lisa) Lee, Charles Gordon, Olivia Shy, and Harold A. Sackeim; **Project administration:** Charles R. Conway, Mark T. Bunker, and Quyen Tran; **Resources:** Charles R. Conway, Scott T. Aaronson, A. John Rush, Ying-Chieh (Lisa) Lee, Charles Gordon, Olivia Shy, Mark T. Bunker, Patricio Riva-Posse, Mark S. George, John Zajecka, Ziad Nahas, David L. Dunner, João L. de Quevedo, Martijn Figee, Brian J. Mickey, Rebecca M. Allen, Kevin Reeves, Donald Bohnenkamp, Christopher L. Kriedt, Vasilis C. Hristidis, Matthew Macaluso, Walter Duffy, Yvette Sheline, Gustavo Alva, Cristina Cusin, Jeffrey I. Bennett, Quyen Tran, and Harold A. Sackeim; **Software:** Ying-Chieh (Lisa) Lee, Charles Gordon, Olivia Shy, and Quyen Tran; **Supervision:** Charles R. Conway, Scott T. Aaronson, A. John Rush, and Harold A. Sackeim; **Validation:** Charles R. Conway, Scott T. Aaronson, A. John Rush, Ying-Chieh (Lisa) Lee, Charles Gordon, Olivia Shy, Mark T. Bunker, Patricio Riva-Posse, Mark S. George, John Zajecka, Ziad Nahas, David L. Dunner, João L. de Quevedo, Martijn Figee, Brian J. Mickey, Rebecca M. Allen, Kevin Reeves, Donald Bohnenkamp, Christopher L. Kriedt, Vasilis C. Hristidis, Charles F. Zorumski, Matthew Macaluso, Walter Duffy, Yvette Sheline, Gustavo Alva, Cristina Cusin, Jeffrey I. Bennett, Quyen Tran, Roger S. McIntyre, Richard Hamish McAllister-Williams, and Harold A. Sackeim; **Visualization:** Charles R. Conway, Scott T. Aaronson, A. John Rush, Ying-Chieh (Lisa) Lee, Charles Gordon, Olivia Shy, and Harold A. Sackeim; **Writing - original draft:** Charles R. Conway, Scott T. Aaronson, A. John Rush, Ying-Chieh (Lisa) Lee, Charles Gordon, Olivia Shy, Mark T. Bunker, Patricio Riva-Posse, João L. de Quevedo, Charles F. Zorumski, Quyen Tran, and Harold A. Sackeim; **Writing - review & editing:** Charles R. Conway, Scott T. Aaronson, A. John Rush, Ying-Chieh (Lisa) Lee, Charles Gordon, Olivia Shy, Mark T. Bunker, Patricio Riva-Posse, Mark S. George, John Zajecka, Ziad Nahas, David L. Dunner, João L. de Quevedo, Martijn Figee, Brian J. Mickey, Rebecca M. Allen, Kevin Reeves, Donald Bohnenkamp, Christopher L. Kriedt, Vasilis C. Hristidis, Charles F. Zorumski, Matthew Macaluso, Walter Duffy, Yvette Sheline, Gustavo Alva, Cristina Cusin, Jeffrey I. Bennett, Quyen Tran, Roger S. McIntyre, Richard Hamish McAllister-Williams, and Harold A. Sackeim.

## Data Availability Statement

The statistical analyses that are reported in the paper will be shared on reasonable request to the corresponding author. The raw data are not publicly available.

