## Supplemental Materials for "The time course, extent, and durability of improvement in depressive symptoms, daily function, and quality of life with three years of vagus nerve stimulation in markedly treatment-resistant major depression: A RECOVER study report"

**Table S1**. Demographic, clinical, and treatment history characteristics of the Early-Active and Delayed-Active groups

| **Variable** | **Total**  **(N=436)** | **Early-Active**  **(N=221)** | **Delayed-Active  (N=215)** | ***P* Value** |
| --- | --- | --- | --- | --- |
| **Demographics** |  |  |  |  |
| Age, yr, mean (SD) | 55.1 (14.2) | 55.4 (14.3) | 54.7 (14.1) | 0.610 |
| Sex, N (%) |  |  |  | 0.188 |
| Female | 287 (65.8) | 152 (68.8) | 135 (62.8) |  |
| Male | 149 (34.2) | 69 (31.2) | 80 (37.2) |  |
| Employed, N (%) | 112 (25.7) | 61 (27.6) | 51 (23.7) | 0.384 |
| **Baseline Clinical Presentation** |  |  |  |  |
| Single-episode vs recurrent MDD, N (%) |  |  |  | 0.348 |
| Single-episode MDD | 201 (46.1) | 97 (43.9) | 104 (48.4) |  |
| Recurrent MDD | 235 (53.9) | 124 (56.1) | 111 (51.6) |  |
| MADRS total score (0–60), mean (SD) | 34.2 (4.7) | 34.1 (4.8) | 34.2 (4.6) | 0.788 |
| CGI-S score (1–7), mean (SD) | 5.1 (0.7) | 5.1 (0.7) | 5.1 (0.8) | 0.927 |
| QIDS-C total score (0–27), mean (SD) | 17.0 (3.1) | 17.2 (3.1) | 16.8 (3.2) | 0.147 |
| QIDS-SR total score (0–27), mean (SD) | 17.5 (3.6) | 17.4 (3.7) | 17.6 (3.4) | 0.436 |
| **Function and Quality of Life Assessments** |  |  |  |  |
| WPAI item 6, mean (SD) | 7.2 (2.0) | 7.2 (2.0) | 7.3 (2.0) | 0.490 |
| Q-LES-Q total score (14–70), mean (SD) | 34.0 (7.0) | 34.2 (7.3) | 33.9 (6.7) | 0.658 |
| Mini-Q-LES-Q total score (7–35), mean (SD) | 15.2 (4.0) | 15.0 (4.1) | 15.3 (3.8) | 0.427 |
| **Mood Disorder History, mean (SD)** |  |  |  |  |
| Age at onset of first symptoms of depression, years | 21.0 (12.2) | 21.0 (12.1) | 20.9 (12.4) | 0.950 |
| Age at definitive diagnosis of any major mood disorder, years | 27.3 (12.5) | 27.3 (12.7) | 27.3 (12.2) | 0.945 |
| Duration of current MDE, years | 17.7 (15.4) | 17.0 (15.2) | 18.4 (15.7) | 0.353 |
| Duration of lifetime MDEs, years^a^ | 29.2 (16.1) | 28.8 (16.5) | 29.5 (15.8) | 0.663 |
| Percentage of lifetime in MDE^b^ | 53.1 (24.5) | 52.1 (24.4) | 54.2 (24.5) | 0.370 |
| Lifetime number of prior hospital admissions for mood disorder | 2.2 (4.0) | 2.3 (4.2) | 2.0 (3.8) | 0.462 |
| Lifetime history of suicide attempts, n (%) | 0.9 (1.8) | 0.9 (1.4) | 0.9 (2.1) | 0.586 |
| **Antidepressant Treatments, Lifetime** |  |  |  |  |
| Lifetime number of inadequate responses to antidepressant treatments, mean (SD)^c^ | 13.3 (8.4) | 13.4 (9.1) | 13.3 (7.6) | 0.873 |
| Lifetime number of inadequate responses to antidepressant pharmacotherapies, mean (SD) | 11.2 (7.3) | 11.4 (8.5) | 11.0 (5.8) | 0.602 |
| Lifetime history of ECT, N (%) |  |  |  | 0.852 |
| Responded | 53 (12.2) | 25 (11.3) | 28 (13.0) |  |
| Did not respond | 153 (35.1) | 79 (35.7) | 74 (34.4) |  |
| **Antidepressant Treatments in Current MDE** |  |  |  |  |
| Any pharmacotherapy, N (%) | 436 (100) | 221 (100) | 215 (100) | NC |
| Esketamine | 107 (24.5) | 49 (22.2) | 58 (27.0) | 0.244 |
| Any psychotherapy, N (%) | 322 (73.9) | 167 (75.6) | 155 (72.1) | 0.409 |
| Any neuromodulation, N (%) | 292 (67.0) | 148 (67.0) | 144 (67.0) | 0.999 |
| TMS | 220 (50.5) | 116 (52.5) | 104 (48.4) | 0.390 |
| ECT | 168 (38.5) | 84 (38.0) | 84 (39.1) | 0.820 |

^a^Duration of lifetime MDEs is the number of years of previous and current major depressive episodes. If both previous and current episodes were missing, then duration of lifetime episodes was considered as missing. If either of the two was not missing, then duration of lifetime episodes was the sum of the non-missing number. The maximum duration of estimated lifetime episodes in years was set as age at baseline. If the number of hospital admissions or of lifetime episodes was more than 10, the number was capped at 10.

^b^Percentage of lifetime in MDE was calculated as “duration of lifetime MDEs” divided by “age at baseline.”

^c^Including pharmacotherapies, ECT, TMS, and psychotherapies.

Abbreviations: CGI-S, Clinical Global Impression–Severity; ECT, electroconvulsive therapy; MADRS, Montgomery-Åsberg Depression Rating Scale; MDD, major depressive disorder; MDE, major depressive episode; Mini-Q-LES-Q, 7-item subset of the Quality of Life Enjoyment and Satisfaction Questionnaire; NC, not calculable; Q-LES-Q, Quality of Life Enjoyment and Satisfaction Questionnaire; QIDS-C, Quick Inventory of Depressive Symptomatology–Clinician; QIDS-SR, Quick Inventory of Depressive Symptomatology–Self-Report; TAU, treatment as usual; TMS, transcranial magnetic stimulation; VNS, vagus nerve stimulation; WPAI, Work Productivity and Activity Impairment Questionnaire.

**Table S2.** Sample size at each time point through 36 months in **A.** Early-Active and **B.** Delayed-Active groups. Sample size is reportedly separately for both all randomized subjects in the mITT population and the subset of subjects who had an assessment at Month 12

A. **Early-Active Group**

|  |  | **Month** | | | | | | | |
| --- | --- | --- | --- | --- | --- | --- | --- | --- | --- |
| **N (% available)** | **Baseline** | **3** | **6** | **9** | **12** | **18** | **24** | **30** | **36** |
| Randomized | | | | | | | | | |
| **MADRS** | 249  (100) | 225 (90.4) | 218 (87.6) | 214 (85.9) | 221 (88.8) | 208 (83.5) | 186 (74.7) | 177 (71.1) | 164 (65.9) |
| **QIDS-C** | 249  (100) | 225 (90.4) | 218 (87.6) | 214 (85.9) | 221 (88.8) | 208 (83.5) | 186 (74.7) | 177 (71.1) | 164 (65.9) |
| **QIDS-SR** | 249  (100) | 225 (90.4) | 217 (87.1) | 211 (84.7) | 220 (88.4) | 209 (83.9) | 187 (75.1) | 178 (71.5) | 166 (66.7) |
| **CGI-I** | 249  (100) | 227 (91.2) | 217 (87.1) | 214 (85.9) | 221 (88.8) | 209 (83.9) | 190 (76.3) | 181 (72.7) | 167 (67.1) |
| **WPAI Item 6** | 248 (99.6) | 221 (88.8) | 214 (85.9) | 207 (83.1) | 217 (87.1) | 206 (82.7) | 183 (73.5) | 177 (71.1) | 163 (65.5) |
| **Mini-Q-LES-Q** | 242 (97.2) | 215 (86.3) | 210 (84.3) | 204 (81.9) | 212 (85.1) | 203 (81.5) | 181 (72.7) | 174 (69.9) | 160 (64.3) |
| **WHODAS** | 249 (100) | 224 (90.0) | 217 (87.1) | 211 (84.7) | 220 (88.4) | 209 (83.9) | 187 (75.1) | 179 (71.9) | 166 (66.7) |
| **EQ-5D-5L VAS** | 249 (100) | 223 (89.6) | 215 (86.3) | 208 (83.5) | 217 (87.1) | 209 (83.9) | 185 (74.3) | 178 (71.5) | 165 (66.3) |
| Randomized with M12 Assessment | | | | | | | | | |
| **MADRS** | - | - | - | - | 221 (100) | 208 (94.1) | 186 (84.2) | 177 (80.1) | 164 (74.2) |
| **QIDS-C** | - | - | - | - | 221 (100) | 208 (94.1) | 186 (84.2) | 177 (80.1) | 164 (74.2) |
| **QIDS-SR** | - | - | - | - | 220 (100) | 208 (94.5) | 186 (84.5) | 177 (80.5) | 165 (75.0) |
| **CGI-I** | - | - | - | - | 221 (100) | 209 (94.6) | 190 (86.0) | 181 (81.9) | 167 (75.6) |
| **WPAI Item 6** | - | - | - | - | 217 (100) | 203 (93.5) | 180 (82.9) | 174 (80.2) | 160 (73.7) |
| **Mini-Q-LES-Q** | - | - | - | - | 212 (100) | 200 (94.3) | 178 (84.0) | 171 (80.7) | 157 (74.1) |
| **WHODAS** | - | - | - | - | 220 (100) | 208 (94.5) | 186 (84.5) | 178 (80.9) | 165 (75.0) |
| **EQ-5D-5L VAS** | - | - | - | - | 217 (100) | 205 (94.5) | 181 (83.4) | 174 (80.2) | 161 (74.2) |

B. **Delayed-Active Group**

|  |  | **Month** | | | | | | | |
| --- | --- | --- | --- | --- | --- | --- | --- | --- | --- |
| **N (% available)** | **Baseline** | **3** | **6** | **9** | **12** | **18** | **24** | **30** | **36** |
| Randomized | | | | | | | | | |
| **MADRS** | 244  (100) | 221 (90.6) | 213 (87.3) | 209 (85.7) | 215 (88.1) | 196 (80.3) | 186 (76.2) | 175 (71.7) | 167 (68.4) |
| **QIDS-C** | 244  (100) | 221 (90.6) | 213 (87.3) | 209 (85.7) | 214 (87.7) | 196 (80.3) | 186 (76.2) | 175 (71.7) | 167 (68.4) |
| **QIDS-SR** | 242 (99.2) | 218 (89.3) | 209 (85.7) | 207 (84.8) | 212 (86.9) | 193 (79.1) | 184 (75.4) | 170 (69.7) | 166 (68.0) |
| **CGI-I** | 244  (100) | 222 (91.0) | 213 (87.3) | 210 (86.1) | 214 (87.7) | 195 (79.9) | 187 (76.6) | 178 (73.0) | 169 (69.3) |
| **WPAI Item 6** | 242 (99.2) | 212 (86.9) | 201 (82.4) | 206 (84.4) | 211 (86.5) | 191 (78.3) | 183 (75.0) | 168 (68.9) | 166 (68.0) |
| **Mini-Q-LES-Q** | 238 (97.5) | 211 (86.5) | 204 (83.6) | 201 (82.4) | 208 (85.2) | 189 (77.5) | 180 (73.8) | 168 (68.9) | 162 (66.4) |
| **WHODAS** | 242 (99.2) | 217 (88.9) | 207 (84.8) | 206 (84.4) | 211 (86.5) | 193 (79.1) | 184 (75.4) | 170 (69.7) | 166 (68.0) |
| **EQ-5D-5L VAS** | 242 (99.2) | 215 (88.1) | 202 (82.8) | 207 (84.8) | 211 (86.5) | 192 (78.7) | 183 (75.0) | 168 (68.9) | 166 (68.0) |
| Randomized with M12 Assessment | | | | | | | | | |
| **MADRS** | - | - | - | - | 215 (100) | 196 (91.2) | 186 (86.5) | 175 (81.4) | 167 (77.7) |
| **QIDS-C** | - | - | - | - | 214 (100) | 195 (91.1) | 185 (86.4) | 175 (81.8) | 166 (77.6) |
| **QIDS-SR** | - | - | - | - | 212 (100) | 192 (90.6) | 183 (86.3) | 170 (80.2) | 166 (78.3) |
| **CGI-I** | - | - | - | - | 214 (100) | 194 (90.7) | 186 (86.9) | 177 (82.7) | 168 (78.5) |
| **WPAI Item 6** | - | - | - | - | 211 (100) | 189 (89.6) | 182 (86.3) | 168 (79.6) | 166 (78.7) |
| **Mini-Q-LES-Q** | - | - | - | - | 208 (100) | 187 (89.9) | 179 (86.1) | 168 (80.8) | 162 (77.9) |
| **WHODAS** | - | - | - | - | 211 (100) | 191 (90.5) | 183 (86.7) | 170 (80.6) | 166 (78.7) |
| **EQ-5D-5L VAS** | - | - | - | - | 211 (100) | 190 (90.0) | 182 (86.3) | 168 (79.6) | 166 (78.7) |

Abbreviations: CGI-I, Clinical Global Impression–Improvement; EQ-5D-5L VAS, EuroQol 5-Dimension 5-Level Visual Analog Scale; MADRS, Montgomery-Åsberg Depression Rating Scale; Mini-Q-LES-Q, 7-item subset of the Quality of Life Enjoyment and Satisfaction Questionnaire; QIDS-C, Quick Inventory of Depressive Symptomatology–Clinician; QIDS-SR, Quick Inventory of Depressive Symptomatology–Self-Report; WHODAS, WHO Disability Assessment Schedule; WPAI, Work Productivity and Activity Impairment Questionnaire.

**Table S3.** Cumulative logit mixed-effects model for exclusive ordinal outcomes (0=no benefit; 1=partial response; 2=response; 3=remission) on four scales

| **Effect** | **Comparison** | **Parameter Estimate (SE)** | **95% CI** | ***P* Value** |
| --- | --- | --- | --- | --- |
| Time | **Monthly change in cumulative log-odds** | **0.079 (0.009)** | **0.062, 0.096** | **<0.0001** |
| Treatment | Early-Active vs Delayed-Active | 0.164 (0.200) | -0.229, 0.557 | 0.413 |
| Treatment × Time | **Difference in monthly treatment slope during the RCT period** | **0.046 (0.012)** | **0.022, 0.069** | **0.0001** |
| Study Phase | **Follow-up vs RCT for Delayed-Active** | **1.114 (0.171)** | **0.778, 1.449** | **<0.0001** |
| Treatment × Study Phase | Change in treatment effect during follow-up relative to RCT | 0.452 (0.237) | -0.012, 0.917 | 0.056 |
| Time × Study Phase | **Change in monthly slope during follow-up relative to RCT** | **-0.055 (0.010)** | **-0.075, -0.034** | **<0.0001** |
| Treatment × Time × Study Phase | **Change in treatment-specific monthly slope during follow-up relative to RCT** | **-0.061 (0.014)** | **-0.089, -0.034** | **<0.0001** |
| Age Group | ≥65 years vs <65 years | 0.328 (0.191) | -0.047, 0.702 | 0.087 |
| Baseline MADRS Category | **≥34 vs <34** | **-0.853 (0.181)** | **-1.208, -0.498** | **<0.0001** |
| Scale | **CGI-I vs QIDS-SR** | **0.311 (0.039)** | **0.234, 0.388** | **<0.0001** |
| Scale | **MADRS vs QIDS-SR** | **-0.522 (0.041)** | **-0.603, -0.442** | **<0.0001** |
| Scale | **QIDS-C vs QIDS-SR** | **-0.198 (0.040)** | **-0.276, -0.119** | **<0.0001** |
| Baseline Severity | **Per 1-SD increase in baseline score** | **0.280 (0.022)** | **0.238, 0.322** | **<0.0001** |

Note: Treatment effects should be interpreted in the context of the treatment-by-time and treatment-by-time-by-study phase interaction terms. Accordingly, the treatment main effect does not represent the overall treatment effect during the study.

Month (time) treated as a continuous variable.

Bolded values represent statistically significant effects.

Abbreviations: CGI-I, Clinical Global Impression–Improvement; CI, confidence interval; MADRS, Montgomery-Åsberg Depression Rating Scale; QIDS-C, Quick Inventory of Depressive Symptomatology–Clinician; QIDS-SR, Quick Inventory of Depressive Symptomatology–Self-Report; RCT, randomized controlled trial; SD, standard deviation.

**Table S4.** Cumulative odds ratio at every visit from Months 3 to 36 comparing Early-Active and Delayed-Active groups from the overall longitudinal analysis

| **Visit** | **Odds Ratio (Lower, Upper)** | ***P* Value** |
| --- | --- | --- |
| Month 3 | 1.35 (0.94, 1.95) | 0.108 |
| Month 4 | 1.41 (0.99, 2.03) | 0.059 |
| Month 5 | **1.48 (1.04, 2.11)** | **0.031** |
| Month 6 | **1.55 (1.09, 2.20)** | **0.015** |
| Month 7 | **1.62 (1.14, 2.30)** | **0.007** |
| Month 8 | **1.70 (1.20, 2.41)** | **0.003** |
| Month 9 | **1.78 (1.25, 2.52)** | **0.001** |
| Month 10 | **1.86 (1.31, 2.65)** | **<0.001** |
| Month 11 | **1.95 (1.36, 2.78)** | **<0.001** |
| Month 12 | **2.04 (1.42, 2.93)** | **<0.001** |
| Overall: Months 3–12 (RCT phase) | **1.66 (1.17, 2.35)** | **0.005** |
| Month 18 | 1.39 (0.95, 2.04) | 0.088 |
| Month 24 | 1.27 (0.88, 1.82) | 0.197 |
| Month 30 | 1.15 (0.80, 1.66) | 0.441 |
| Month 36 | 1.05 (0.71, 1.54) | 0.809 |
| Overall: Months 18-36 | 1.21 (0.85, 1.73) | 0.299 |

OR >1 favors the Early-Active group.

Bolded values represent statistical significance.

Abbreviations: OR, odds ratio; RCT, randomized controlled trial.

**Table S5.** Summary of total scores presented as mean (SD) through 36 months in both A. Early-Active and B. Delayed-Active groups

A. **Early-Active Group**

|  |  | **Month** | | | | | | | |
| --- | --- | --- | --- | --- | --- | --- | --- | --- | --- |
| **Mean (SD) Total Score** | **Baseline** | **3** | **6** | **9** | **12** | **18** | **24** | **30** | **36** |
| **MADRS** | 34.1 (4.8) | 27.1 (9.4) | 25.5 (11.1) | 25.4 (10.7) | 23.2 (11.4) | 22.2 (11.5) | 20.8 (11.1) | 20.5 (11.5) | 20.4 (12.2) |
| **QIDS-C** | 17.2 (3.1) | 13.4 (5.1) | 12.3 (5.6) | 12.0 (5.3) | 11.0 (5.5) | 10.7 (5.6) | 10.1 (5.5) | 9.7 (5.4) | 9.7 (5.6) |
| **QIDS-SR** | 17.4 (3.7) | 13.8 (5.1) | 12.1 (5.5) | 12.0 (5.9) | 11.0 (6.0) | 10.9 (5.7) | 10.6 (5.7) | 10.4 (5.6) | 10.5 (5.8) |
| **CGI-I** | 5.1 (0.7) | 3.4 (1.1) | 3.1 (1.2) | 3.1 (1.2) | 2.8 (1.2) | 2.7 (1.1) | 2.5 (1.2) | 2.5 (1.2) | 2.5 (1.3) |
| **WPAI Item 6** | 71.8 (19.7) | 57.3 (27.7) | 50.7 (26.9) | 52.2 (28.5) | 48.1 (30.3) | 44.9 (29.8) | 45.6 (29.3) | 42.9 (29.0) | 43.3 (29.6) |
| **Mini-Q-LES-Q** | 28.7 (14.7) | 41.0 (19.6) | 44.7 (20.6) | 44.9 (19.7) | 49 (21.3) | 49.2 (21.9) | 51.7 (20.9) | 52.7 (21.1) | 52.7 (22.9) |
| **WHODAS** | 48.2 (17.7) | 40.3 (19.9) | 37.7 (19.4) | 37.3 (20.1) | 36.0 (20.7) | 34.5 (19.3) | 33.4 (18.6) | 32.5 (19.4) | 33.2 (20.2) |
| **EQ-5D-5L VAS** | 52.9 (21.2) | 56.3 (20.9) | 55.8 (20.2) | 57.6 (19.9) | 60.5 (20.4) | 59.5 (19.9) | 59.6 (19.6) | 59.7 (21.4) | 61.0 (21.1) |

B. **Delayed-Active Group**

|  |  | **Month** | | | | | | | |
| --- | --- | --- | --- | --- | --- | --- | --- | --- | --- |
|  | **Baseline** | **3** | **6** | **9** | **12** | **18** | **24** | **30** | **36** |
| **MADRS** | 34.2 (4.6) | 27.4 (10.1) | 26.6 (9.5) | 25.8 (10.6) | 24.8 (11.9) | 23.3 (10.9) | 23.0 (11.1) | 21.9 (11.7) | 22.1 (11.6) |
| **QIDS-C** | 16.8 (3.2) | 13.5 (5.2) | 13.1 (4.9) | 12.5 (5.2) | 12.2 (5.7) | 11.2 (5.5) | 10.8 (5.4) | 10.4 (5.8) | 10.4 (5.5) |
| **QIDS-SR** | 17.6 (3.4) | 13.3 (5.4) | 13.1 (5.5) | 12.6 (5.8) | 12.1 (6.0) | 11.7 (5.5) | 11.8 (5.6) | 11.3 (6.0) | 10.9 (5.9) |
| **CGI-I** | 5.1 (0.8) | 3.5 (1.0) | 3.4 (1.0) | 3.4 (1.1) | 3.3 (1.1) | 2.9 (1.1) | 2.7 (1.2) | 2.6 (1.3) | 2.7 (1.3) |
| **WPAI Item 6** | 73.3 (20.2) | 57.5 (26.4) | 58.5 (27.1) | 56.3 (27.3) | 56.3 (30.5) | 50.8 (28.4) | 48.3 (29.9) | 49.0 (30.4) | 48.9 (29.7) |
| **Mini-Q-LES-Q** | 29.7 (13.7) | 41.1 (19.3) | 41.2 (18.9) | 42.2 (20.7) | 43.6 (21.3) | 46.9 (20.3) | 46.0 (20.5) | 47.6 (20.5) | 48.4 (20.9) |
| **WHODAS** | 48.4 (15.5) | 40.1 (18.8) | 41.4 (20.0) | 40.2 (19.7) | 39.3 (20.3) | 36.7 (19.9) | 37.9 (19.5) | 36.9 (20.0) | 36.9 (18.9) |
| **EQ-5D-5L VAS** | 54.3 (18.6) | 55.3 (20.5) | 54.0 (20.4) | 56.8 (20.7) | 57.3 (21.2) | 58.0 (20.5) | 58.5 (20.3) | 58.3 (20.6) | 57.6 (22.2) |

Abbreviations: CGI-I, Clinical Global Impression–Improvement; EQ-5D-5L VAS, EuroQol 5-Dimension 5-Level Visual Analog Scale; MADRS, Montgomery-Åsberg Depression Rating Scale; Mini-Q-LES-Q, 7-item subset of the Quality of Life Enjoyment and Satisfaction Questionnaire; QIDS-C, Quick Inventory of Depressive Symptomatology–Clinician; QIDS-SR, Quick Inventory of Depressive Symptomatology–Self-Report; WHODAS, WHO Disability Assessment Schedule; WPAI, Work Productivity and Activity Impairment Questionnaire.

**Table S6.** Cumulative logit mixed-effects model assessing within-group change in extent of categorical benefit for each time interval for the MADRS, QIDS-C, QIDS-SR, CGI-I, WPAI item 6, and Mini-Q-LES-Q

| **Outcome Measure** | **Comparison** | **Odds Ratio (95% CI); *P*** | |
| --- | --- | --- | --- |
|  |  | **Early-Active** | **Delayed-Active** |
| **MADRS** | Month 12 to 24 | **1.6 (1.1, 2.5); 0.023** | **1.8 (1.1, 2.8); 0.011** |
|  | Month 24 to 36 | 1.2 (0.7, 1.8); 0.525 | 1.3 (0.8, 2.0); 0.307 |
|  | Month 12 to 36 | **1.9 (1.2, 3.0); 0.004** | **2.3 (1.4, 3.6); <0.001** |
| **QIDS-C** | Month 12 to 24 | 1.5 (1.0, 2.2); 0.056 | **2.3 (1.5, 3.5); <0.001** |
|  | Month 24 to 36 | 0.9 (0.6, 1.4); 0.699 | 1.3 (0.9, 2.0); 0.220 |
|  | Month 12 to 36 | 1.4 (0.9, 2.1); 0.150 | **3.0 (2.0, 4.7); <0.001** |
| **QIDS-SR** | Month 12 to 24 | 0.9 (0.6, 1.3); 0.553 | 1.3 (0.9, 2.0); 0.207 |
|  | Month 24 to 36 | 0.9 (0.6, 1.5); 0.762 | 1.4 (0.9, 2.1); 0.151 |
|  | Month 12 to 36 | 0.8 (0.5, 1.3); 0.374 | **1.8 (1.2, 2.7); 0.007** |
| **CGI-I** | Month 12 to 24 | **1.6 (1.1, 2.3); 0.010** | **3.3 (2.3, 4.8); <0.001** |
|  | Month 24 to 36 | 1.0 (0.7, 1.4); 0.935 | 1.1 (0.7, 1.6); 0.752 |
|  | Month 12 to 36 | **1.6 (1.1, 2.3); 0.016** | **3.5 (2.4, 5.2); <0.001** |
| **WPAI Item 6** | Month 12 to 24 | 1.2 (0.8, 2.0); 0.381 | **1.8 (1.2, 3.0); 0.010** |
|  | Month 24 to 36 | 1.2 (0.7, 2.1); 0.445 | 1.0 (0.6, 1.7); 0.990 |
|  | Month 12 to 36 | 1.5 (0.9, 2.5); 0.103 | **1.9 (1.1, 3.0); 0.012** |
| **Mini-Q-LES-Q** | Month 12 to 24 | 1.3 (0.8, 2.1); 0.346 | 1.5 (0.9, 2.5); 0.092 |
|  | Month 24 to 36 | 0.9 (0.5, 1.5); 0.581 | 1.3 (0.8, 2.3); 0.280 |
|  | Month 12 to 36 | 1.1 (0.6, 1.9); 0.744 | **2.0 (1.2, 3.4); 0.006** |

OR >1 = greater odds of improved status at the later visit. Contrasts are within each arm over 36 months, not between arms.

Bolded data show significant values (*P*<0.05 unadjusted)

Abbreviations: CGI-I, Clinical Global Impression–Improvement; CI, confidence interval; MADRS, Montgomery-Åsberg Depression Rating Scale; Mini-Q-LES-Q, 7-item subset of the Quality of Life Enjoyment and Satisfaction Questionnaire; OR, odds ratio; QIDS-C, Quick Inventory of Depressive Symptomatology–Clinician; QIDS-SR, Quick Inventory of Depressive Symptomatology–Self-Report; WPAI, Work Productivity and Activity Impairment Questionnaire.

**Table S7.** Mixed-effects model assessing within-group change in total score for all measures

| **Outcome Measure (Direction of improvement**) | **Comparison** | **Change in Total Score (95% CI); *P**** | |
| --- | --- | --- | --- |
|  |  | **Early-Active** | **Delayed-Active** |
| **MADRS (↓)** | Month 12 to 24 | **-2.0 (-3.3, -0.7); 0.003** | **-1.9 (-3.2, -0.6); 0.004** |
|  | Month 24 to 36 | 0.0 (-1.4, 1.4); 0.993 | -1.1 (-2.4, 0.3); 0.133 |
|  | Month 12 to 36 | **-2.0 (-3.3, -0.6); 0.004** | **-2.9 (-4.2, -1.6); <0.001** |
| **QIDS-C (↓)** | Month 12 to 24 | **-0.8 (-1.4, -0.1); 0.022** | **-1.4 (-2.1, -0.7); <0.001** |
|  | Month 24 to 36 | -0.2 (-0.9, 0.6); 0.654 | -0.5 (-1.2, 0.3); 0.202 |
|  | Month 12 to 36 | **-0.9 (-1.6, -0.3); 0.008** | **-1.9 (-2.5, -1.2); <0.001** |
| **QIDS-SR (↓)** | Month 12 to 24 | -0.2 (-0.8, 0.5); 0.602 | -0.3 (-1.0, 0.3); 0.345 |
|  | Month 24 to 36 | 0.2 (-0.5, 0.9); 0.559 | **-0.9 (-1.6, -0.2); 0.011** |
|  | Month 12 to 36 | 0.0 (-0.6, 0.7); 0.919 | **-1.2 (-1.9, -0.5); <0.001** |
| **CGI-I (↓)** | Month 12 to 24 | **-0.2 (-0.4, -0.1); 0.003** | **-0.6 (-0.8, -0.5); <0.001** |
|  | Month 24 to 36 | 0.0 (-0.2, 0.2); 0.955 | 0.0 (-0.2, 0.1); 0.711 |
|  | Month 12 to 36 | **-0.2 (-0.4, -0.1); 0.004** | **-0.7 (-0.8, -0.5); <0.001** |
| **WPAI Item 6 (↓)** | Month 12 to 24 | -0.2 (-0.6, 0.2); 0.332 | **-0.8 (-1.2, -0.4); <0.001** |
|  | Month 24 to 36 | 0.0 (-0.5, 0.4); 0.929 | 0.1 (-0.3, 0.5); 0.643 |
|  | Month 12 to 36 | -0.2 (-0.6, 0.2); 0.304 | **-0.7 (-1.1, -0.3); <0.001** |
| **Mini-Q-LES-Q (↑)** | Month 12 to 24 | 0.7 (-0.1, 1.4); 0.084 | 0.7 (-0.0, 1.5); 0.063 |
|  | Month 24 to 36 | -0.1 (-0.9, 0.7); 0.893 | 0.7 (-0.1, 1.5); 0.100 |
|  | Month 12 to 36 | 0.6 (-0.2, 1.4); 0.128 | **1.4 (0.6, 2.1); <0.001** |
| **WHODAS (↓)** | Month 12 to 24 | **-2.2 (-4.3, -0.1); 0.045** | -1.4 (-3.5, 0.8); 0.207 |
|  | Month 24 to 36 | 0.7 (-1.6, 3.0); 0.540 | -1.0 (-3.2, 1.3); 0.397 |
|  | Month 12 to 36 | -1.4 (-3.6, 0.7); 0.194 | **-2.3 (-4.5, -0.2); 0.036** |
| **EQ-5D-5L VAS (↑)** | Month 12 to 24 | -1.3 (-4.1, 1.5); 0.372 | 1.5 (-1.4, 4.3); 0.310 |
|  | Month 24 to 36 | 0.8 (-2.2, 3.8); 0.609 | -1.1 (-4.1, 1.9); 0.477 |
|  | Month 12 to 36 | -0.5 (-3.4, 2.4); 0.742 | 0.4 (-2.5, 3.3); 0.800 |

Bolded data show significant values.

Abbreviations: CGI-I, Clinical Global Impression–Improvement; CI, confidence interval; EQ-5D-5L VAS, EuroQol 5-Dimension 5-Level Visual Analog Scale; MADRS, Montgomery-Åsberg Depression Rating Scale; Mini-Q-LES-Q, 7-item subset of the Quality of Life Enjoyment and Satisfaction Questionnaire; QIDS-C, Quick Inventory of Depressive Symptomatology–Clinician; QIDS-SR, Quick Inventory of Depressive Symptomatology–Self-Report; WHODAS, WHO Disability Assessment Schedule; WPAI, Work Productivity and Activity Impairment Questionnaire.

**Table S8.** Proportion of participants achieving specific categorical outcomes on individual measures at Month 12, 24, and 36 assessments for both the Early-Active and Delayed-Active groups and within-group comparisons of change in dependent proportions. Data presented are based on inclusive outcome definitions

A. **Early-Active Group**

|  | **Months** | | | **%Change (95% CI); *P*** | | |
| --- | --- | --- | --- | --- | --- | --- |
| **Outcome Classification** | **12** | **24** | **36** | **Months  12 to 24** | **Months  24 to 36** | **Months  12 to 36** |
|  | **MADRS** | | | | | |
| Partial Response | 43.9% | 53.2% | 56.7% | **9.1%  (2.2%, 16.1%);  0.011** | 0.0%  (-7.7%, 7.7%);  1.000 | **8.5%  (0.2%, 16.9%);  0.048** |
| Response | 28.1% | 33.3% | 39.0% | 4.3%  (-2.0%, 10.6%); 0.182 | 2.6%  (-5.0%, 10.1%);  0.505 | **8.5%  (0.5%, 16.5%);  0.039** |
| Remission | 15.8% | 21.0% | 26.8% | 5.4%  (-0.3%, 11.1%);  0.068 | 3.8%  (-2.8%, 10.5%);  0.257 | **9.8%  (2.7%, 16.8%);  0.008** |
|  | **QIDS-C** | | | | | |
| Partial Response | 56.1% | 61.8% | 61.6% | 5.9%  (-1.6%, 13.4%);  0.123 | -3.8%  (-11.6%, 3.9%);  0.330 | 1.2%  (-7.2%, 9.7%);  0.777 |
| Response | 33.9% | 40.9% | 44.5% | **7.5%  (1.0%, 14.1%);  0.027** | -0.6%  (-8.7%, 7.4%);  0.876 | **8.5%  (0.5%, 16.5%);  0.039** |
| Remission | 19.0% | 24.2% | 25.6% | 4.8%  (-1.5%, 11.2%);  0.139 | -2.6%  (-9.7%, 4.5%);  0.480 | 4.9%  (-2.6%, 12.4%);  0.206 |
|  | **QIDS-SR** | | | | | |
| Partial Response | 59.1% | 58.6% | 58.8% | -2.2%  (-9.0%, 4.7%);  0.537 | -5.1%  (-13.6%, 3.4%);  0.238 | -4.8%  (-13.7%, 4.0%);  0.285 |
| Response | 37.3% | 40.9% | 38.8% | 2.7%  (-4.1%, 9.4%);  0.435 | -7.1%  (-15.4%, 1.3%);  0.101 | -1.8%  (-10.0%, 6.3%);  0.662 |
| Remission | 24.1% | 23.1% | 23.6% | -1.6%  (-8.7%, 5.5%);  0.655 | -0.6%  (-6.9%, 5.6%);  0.841 | -3.6%  (-11.3%, 4.0%);  0.355 |
|  | **CGI-I** | | | | | |
| Partial Response | 67.4% | 77.4% | 73.7% | **9.5%  (1.9%, 17.1%);  0.016** | **-8.9%  (-16.0%, -1.8%);  0.016** | 1.8%  (-6.7%, 10.3%);  0.680 |
| Response | 38.9% | 51.6% | 55.1% | **11.1%  (4.2%, 18.0%);  0.002** | -3.2%  (-10.7%, 4.4%);  0.411 | **9.6%  (0.6%, 18.6%) 0.039** |
| Remission | 16.3% | 22.1% | 28.7% | 4.7%  (-0.8%, 10.3%);  0.095 | 1.9%  (-5.6%, 9.4%);  0.622 | **9.0%  (1.2%, 16.7%);  0.025** |
|  | **WPAI Item 6** | | | | | |
| ≥MB | 55.8% | 60.0% | 65.6% | 3.9%  (-3.7%, 11.5%);  0.317 | -1.4%  (-9.7%, 7.0%);  0.752 | 3.8%  (-4.4%, 11.9%);  0.366 |
|  | **Mini-Q-LES-Q** | | | | | |
| ≥MB | 61.8% | 66.3% | 65.0% | 3.4%  (-4.2%, 11.0%);  0.386 | -4.1%  (-12.7%, 4.5%);  0.355 | 0.6%  (-8.1%, 9.4%);  0.886 |

B. **Delayed-Active Group**

|  | **Months** | | | **%Change (95% CI), *P*** | | |
| --- | --- | --- | --- | --- | --- | --- |
| **Outcome Classification** | **12** | **24** | **36** | **Months  12 to 24** | **Months  24 to 36** | **Months 12 to 36** |
|  | **MADRS** | | | | | |
| Partial Response | 36.3% | 48.9% | 47.3% | **12.4%  (5.4%, 19.4%);  <0.001** | -3.1%  (-11.1%, 4.9%);  0.446 | **11.4%  (2.9%, 19.9%);  0.010** |
| Response | 26.0% | 26.9% | 31.1% | 0.5%  (-5.9%, 7.0%);  0.869 | 3.1%  (-4.3%, 10.5%);  0.411 | 6.6%  (-1.4%, 14.6%);  0.109 |
| Remission | 14.9% | 15.1% | 20.4% | 1.1%  (-3.6%, 5.8%);  0.655 | 4.3%  (-1.7%, 10.4%);  0.162 | **8.4%  (2.3%, 14.5%);  0.008** |
|  | **QIDS-C** | | | | | |
| Partial Response | 40.7% | 53.5% | 56.0% | **13.0%  (5.3%, 20.7%);  0.001** | 3.8%  (-4.4%, 11.9%);  0.366 | **16.3%  (7.1%, 25.5%);  <0.001** |
| Response | 24.8% | 35.7% | 38.6% | **11.4%  (4.6%, 18.1%);  0.001** | 2.5%  (-4.6%, 9.6%);  0.493 | **14.5%  (6.2%, 22.7%);  <0.001** |
| Remission | 16.4% | 18.9% | 22.9% | 3.8%  (-2.5%, 10.0%);  0.237 | 1.9%  (-4.9%, 8.7%);  0.590 | **8.4%  (1.7%, 15.2%); 0.016** |
|  | **QIDS-SR** | | | | | |
| Partial Response | 49.1% | 57.9% | 58.4% | **8.7%  (1.3%, 16.2%);  0.024** | 0.0%  (-7.6%, 7.6%);  1.000 | **10.2%  (1.8%, 18.7%);  0.020** |
| Response | 30.2% | 31.7% | 38.0% | 2.2%  (-4.4%, 8.8%);  0.516 | 5.7%  (-2.0%, 13.3%);  0.150 | **9.0%  (2.2%, 15.9%);  0.011** |
| Remission | 17.5% | 15.3% | 21.1% | -1.6%  (-6.8%, 3.5%);  0.532 | 4.4%  (-1.5%, 10.3%); 0144 | 4.8%  (-1.4%, 11.0%);  0.131 |
|  | **CGI-I** | | | | | |
| Partial Response | 44.9% | 73.7% | 73.8% | **28.5%  (20.8%, 36.2%);  <0.001** | -3.7%  (-11.3%, 3.9%);  0.343 | **25.0%  (15.6%, 34.4%);  <0.001** |
| Response | 22.4% | 43.5% | 47.6% | **20.4%  (13.3%, 27.6%);  <0.001** | 0.6% (-6.7%, 8.0%) 0.869 | **22.6%  13.9%, 31.3%);  <0.001** |
| Remission | 6.5% | 18.8% | 22.0% | **12.9%  (7.9%, 18.0%);  <0.001** | 0.6%  (-5.4%, 6.7%);  0.841 | **16.1%  (9.8%, 22.3%);  <0.001** |
|  | **WPAI Item 6** | | | | | |
| ≥MB | 46.9% | 58.2% | 58.4% | **9.9%  (2.4%, 17.4%);  0.011** | 0.0%  (-8.6%, 8.6%);  1.000 | **10.8%  (1.9%, 19.8%);  0.020** |
|  | **Mini-Q-LES-Q** | | | | | |
| ≥MB | 46.2% | 53.6% | 58.6% | 7.3%  (-0.5%, 15.0%);  0.069 | 3.9%  (-3.9%, 11.7%);  0.330 | **13.6%  (4.9%, 22.2%);  0.003** |

Dependent proportions use samples with data at both visits.

Bolded data show significant values.

Abbreviations: CGI-I, Clinical Global Impression–Improvement; CI, confidence interval; MADRS, Montgomery-Åsberg Depression Rating Scale; MB, meaningful benefit; Mini-Q-LES-Q, 7-item subset of the Quality of Life Enjoyment and Satisfaction Questionnaire; QIDS-C, Quick Inventory of Depressive Symptomatology–Clinician; QIDS-SR, Quick Inventory of Depressive Symptomatology–Self-Report; TRD, treatment-resistant depression; VNS, vagus nerve stimulation; WPAI, Work Productivity and Activity Impairment Questionnaire.

**Table S9.** Proportion of participants achieving specific categorical outcomes on individual measures in those who previously failed on an interventional treatment (ECT, TMS, esketamine) for both the Early-Active and Delayed-Active groups. Data presented are based on inclusive outcome definitions

A. Durability of partial response (partial response at Month 12 to partial response at Month 36)

|  | **Early-Active** | | **Delayed-Active** | |
| --- | --- | --- | --- | --- |
| **Outcome Measure** | **Total** | **Intervention**  **Sub-Population** | **Total** | **Intervention**  **Sub-Population** |
| **MADRS** | 77.2% (61/79) | 71.7% (38/53) | 70.0% (42/60) | 70.7% (29/41) |
| **QIDS-C** | 75.8% (75/99) | 72.5% (50/69) | 71.2% (47/66) | 71.7% (33/46) |
| **QIDS-SR** | 69.5% (73/105) | 65.7% (46/70) | 77.5% (62/80) | 76.8% (43/56) |
| **CGI-I** | 79.2% (95/120) | 79.0% (64/81) | 79.3% (65/82) | 78.9% (45/57) |
| **WPAI Item 6^a^** | 80.8% (80/99) | 79.7% (55/69) | 73.4% (58/79) | 67.9% (38/56) |
| **Mini-Q-LES-Q^a^** | 76.2% (77/101) | 74.6% (50/67) | 78.1% (57/73) | 76.6% (36/47) |
| **Median** | **76.7%** | **73.6%** | **75.5%** | **74.2%** |

B. Durability of response (response at Month 12 to response at Month 36)

|  | **Early-Active** | | **Delayed-Active** | |
| --- | --- | --- | --- | --- |
| **Outcome Measure** | **Total** | **Intervention**  **Sub-Population** | **Total** | **Intervention**  **Sub-Population** |
| **MADRS** | 68.0% (34/50) | 58.6% (17/29) | 56.1% (23/41) | 55.6% (15/27) |
| **QIDS-C** | 72.9% (43/59) | 63.9% (23/36) | 65.0% (26/40) | 64.0% (16/25) |
| **QIDS-SR** | 62.7% (42/67) | 60.5% (26/43) | 79.2% (38/48) | 75.9% (22/29) |
| **CGI-I** | 71.1% (54/76) | 68.0% (34/50) | 69.0% (29/42) | 64.5% (20/31) |
| **Median** | **69.6%** | **62.2%** | **67.0%** | **64.3%** |

C. Durability of remission (remission at Month 12 to remission at Month 36)

|  | **Early-Active** | | **Delayed-Active** | |
| --- | --- | --- | --- | --- |
| **Outcome Measure** | **Total** | **Intervention**  **Sub-Population** | **Total** | **Intervention**  **Sub-Population** |
| **MADRS** | 64.3% (18/28) | 62.5% (10/16) | 65.0% (13/20) | 64.3% (9/14) |
| **QIDS-C** | 52.9% (18/34) | 47.6% (10/21) | 58.3% (14/24) | 50.0% (8/16) |
| **QIDS-SR** | 46.7% (21/45) | 44.4% (12/27) | 63.0% (17/27) | 60.0% (9/15) |
| **CGI-I** | 54.5% (18/33) | 60.0% (9/15) | 70.0% (7/10) | 62.5% (5/8) |
| **Median** | **53.7%** | **53.8%** | **64.0%** | **61.3%** |

^a^Partial response equates to meaningful benefit for these outcome measures.

Abbreviations: CGI-I, Clinical Global Impression–Improvement; ECT, electroconvulsive therapy; MADRS, Montgomery-Åsberg Depression Rating Scale; MB, meaningful benefit; Mini-Q-LES-Q, 7-item subset of the Quality of Life Enjoyment and Satisfaction Questionnaire; QIDS-C, Quick Inventory of Depressive Symptomatology–Clinician; QIDS-SR, Quick Inventory of Depressive Symptomatology–Self-Report; TMS, transcranial magnetic stimulation; WPAI, Work Productivity and Activity Impairment Questionnaire.

**Figure S1.** Average outcome score across the MADRS, QIDS-C, QIDS-SR, and CGI-I at each assessment by treatment group with participants scored as 0=no benefit; 1=partial response; 2=response; 3=remission. Data are based on exclusive outcome definitions.

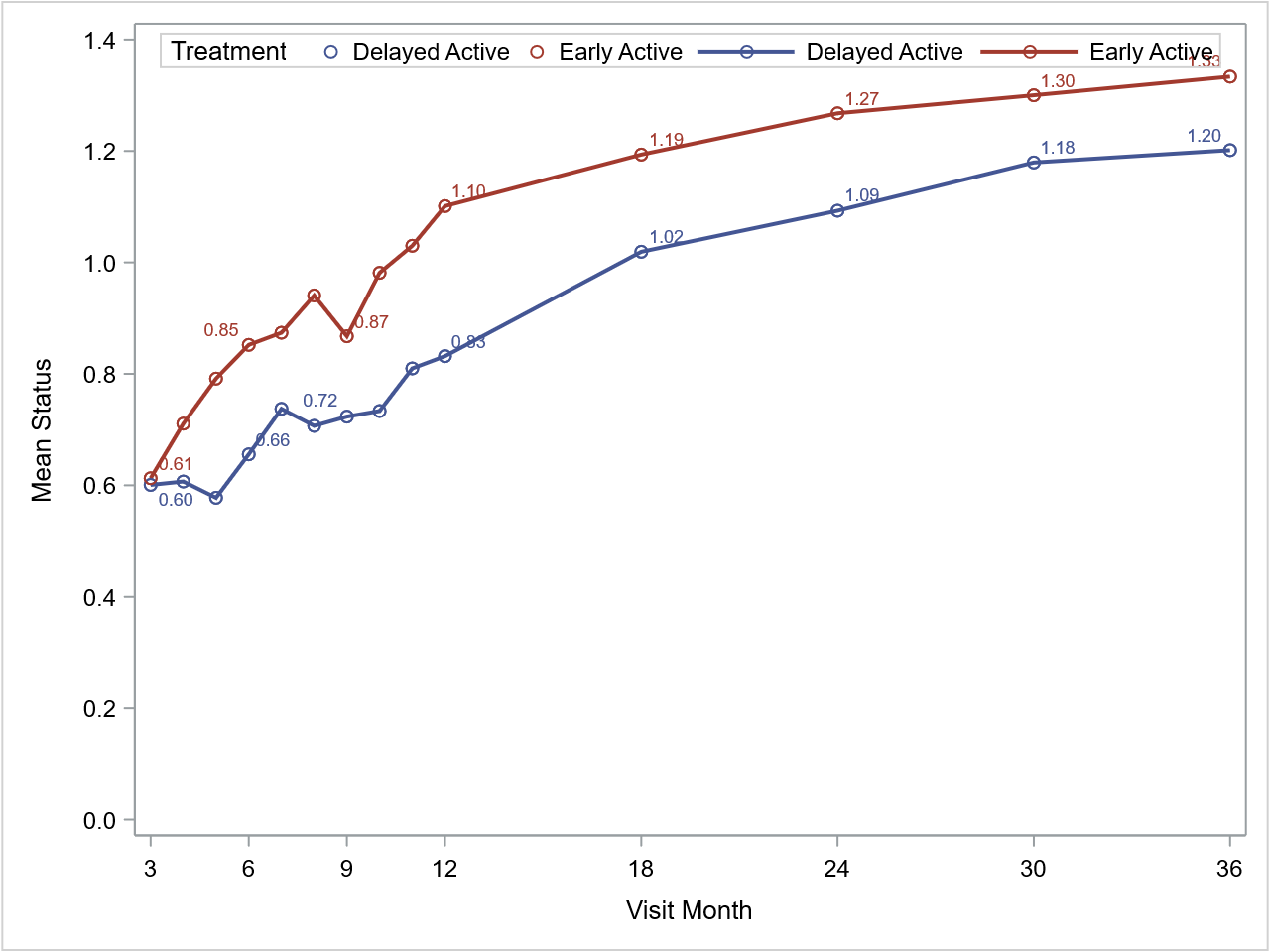

Abbreviations: CGI-I, Clinical Global Impression–Improvement; MADRS, Montgomery-Åsberg Depression Rating Scale; QIDS-C, Quick Inventory of Depressive Symptomatology–Clinician; QIDS-SR, Quick Inventory of Depressive Symptomatology–Self-Report.

**Figure S2**. Alluvial diagrams for change in **A., B.** QIDS-SR; **C., D.** Mini-Q-LES-Q; and **E., F.** WPAI item 6 outcome categories in the Early-Active and Delayed-Active groups from Month 12 to Month 36. Data are based on exclusive outcome definitions.

| A. Early-Active Group: QIDS-SR   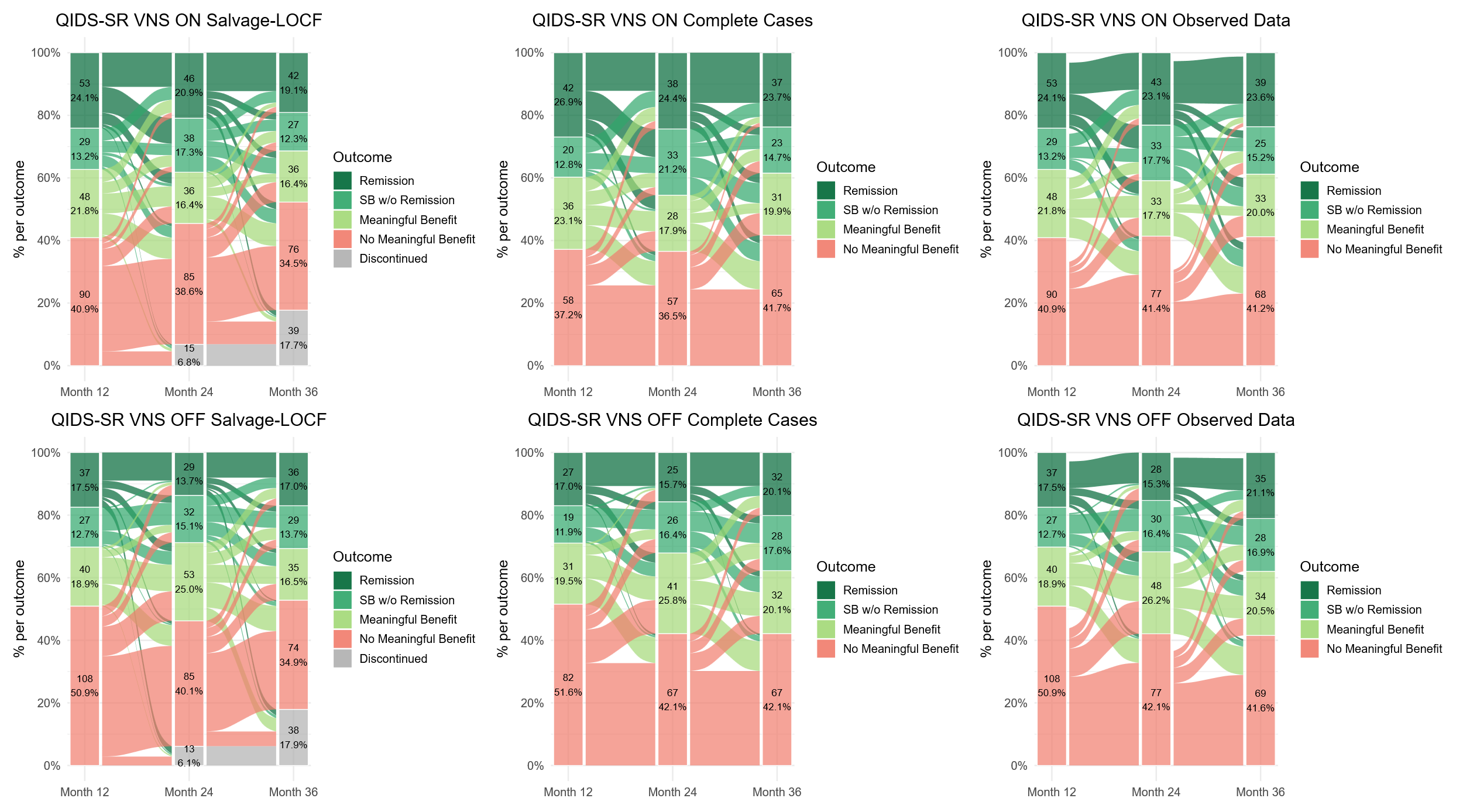 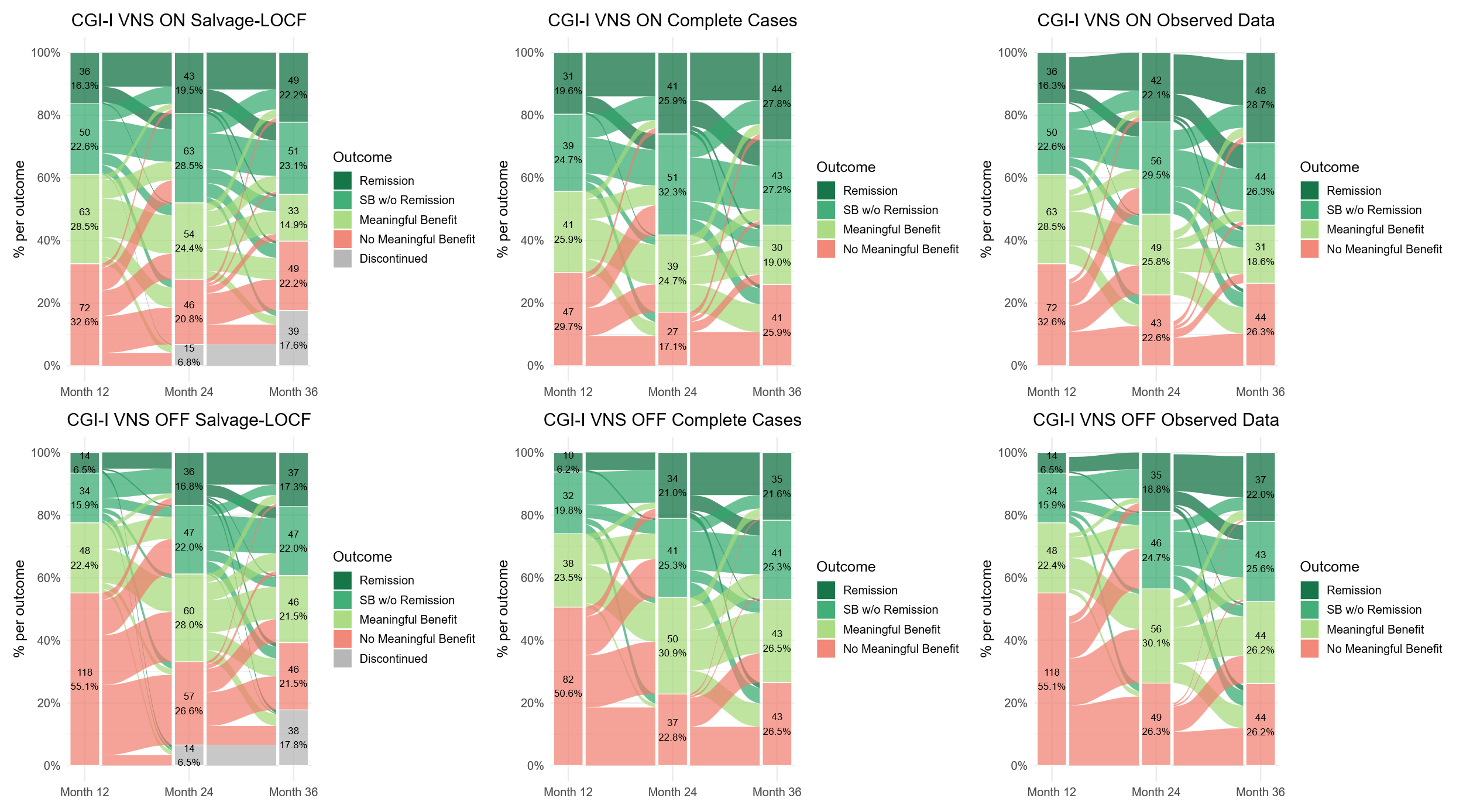 Remission  Response w/o remission  Partial response w/o response  No meaningful benefit | B. Delayed-Active Group: QIDS-SR   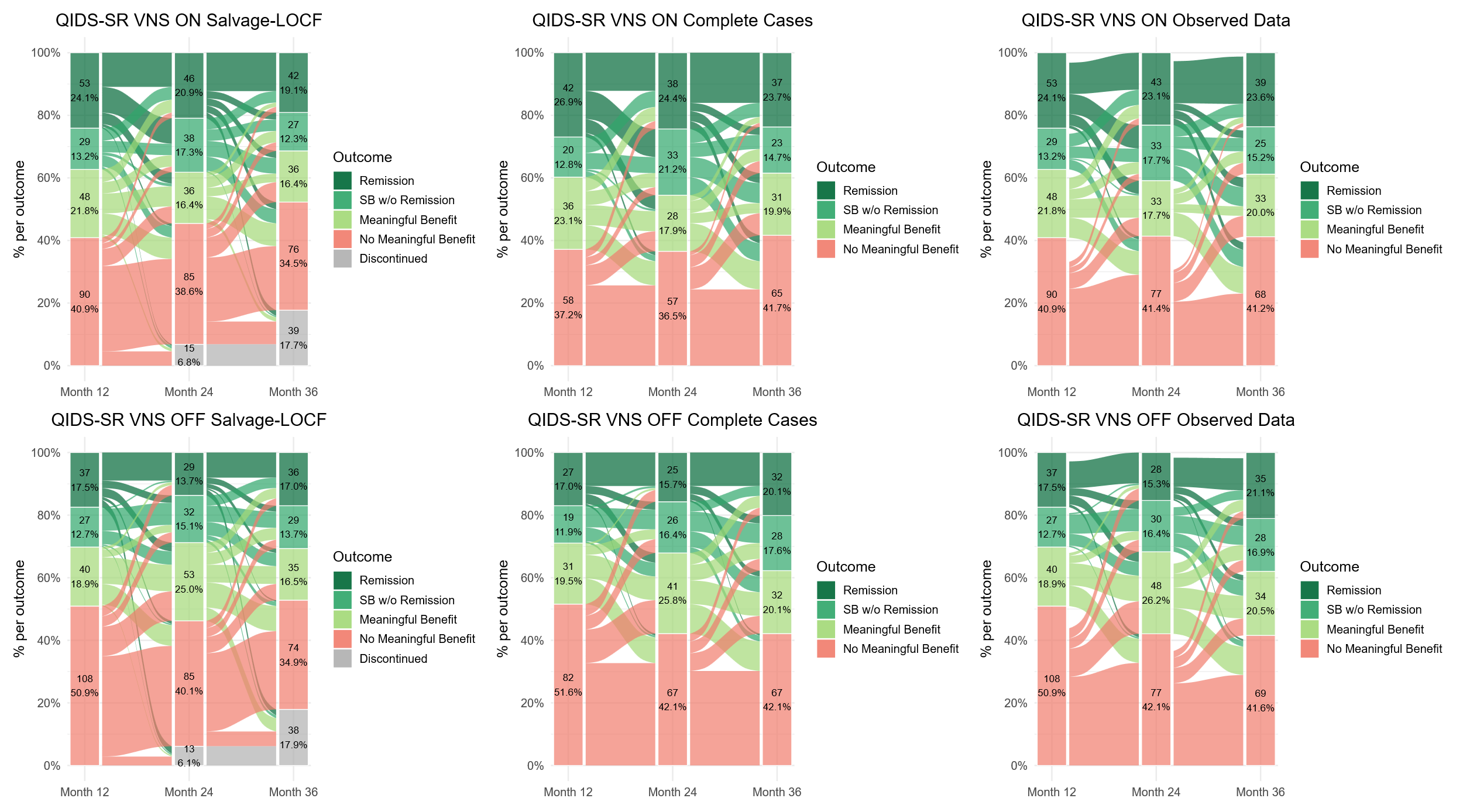 |
| --- | --- |
| C. Early-Active Group: WPAI Item 6   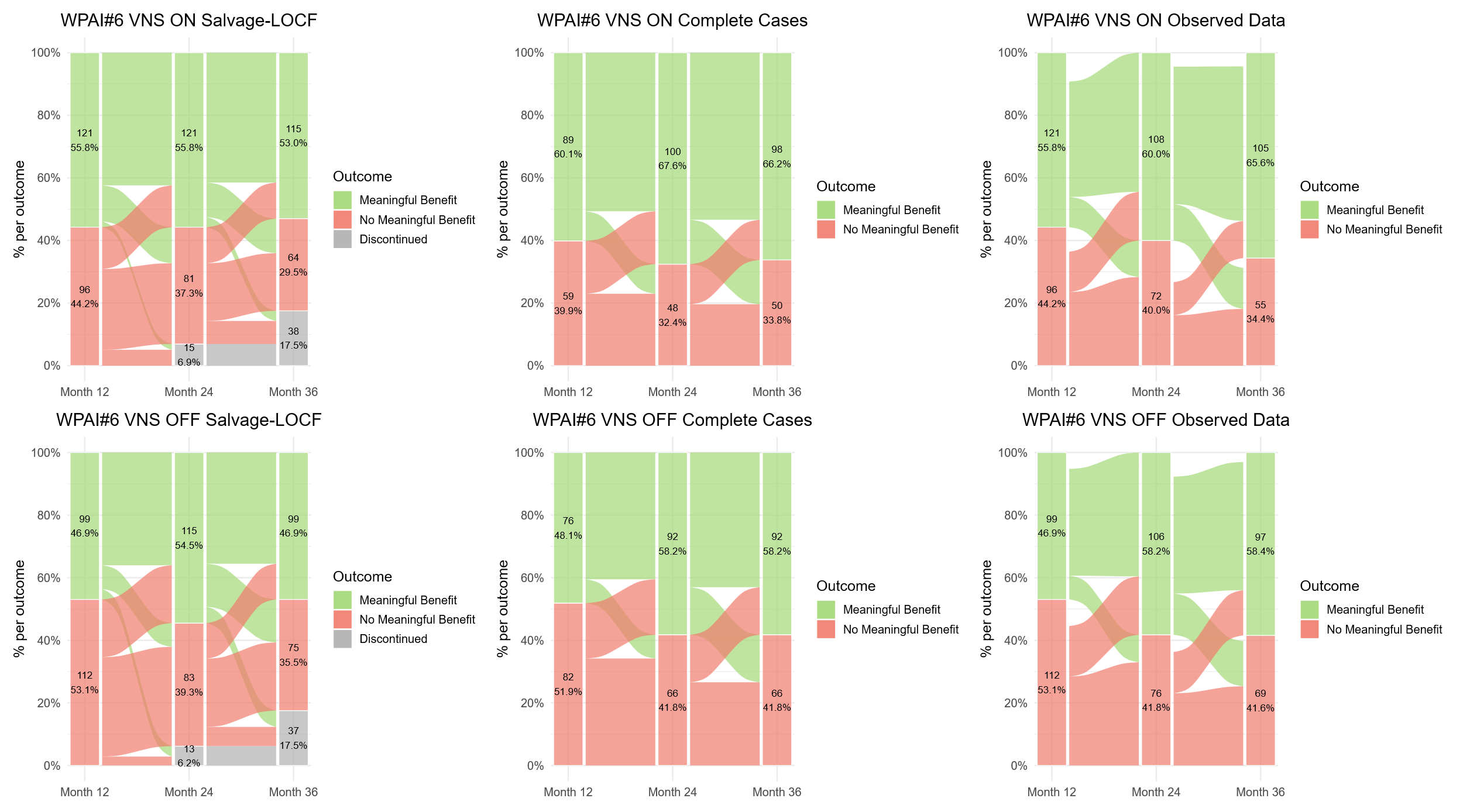 | D. 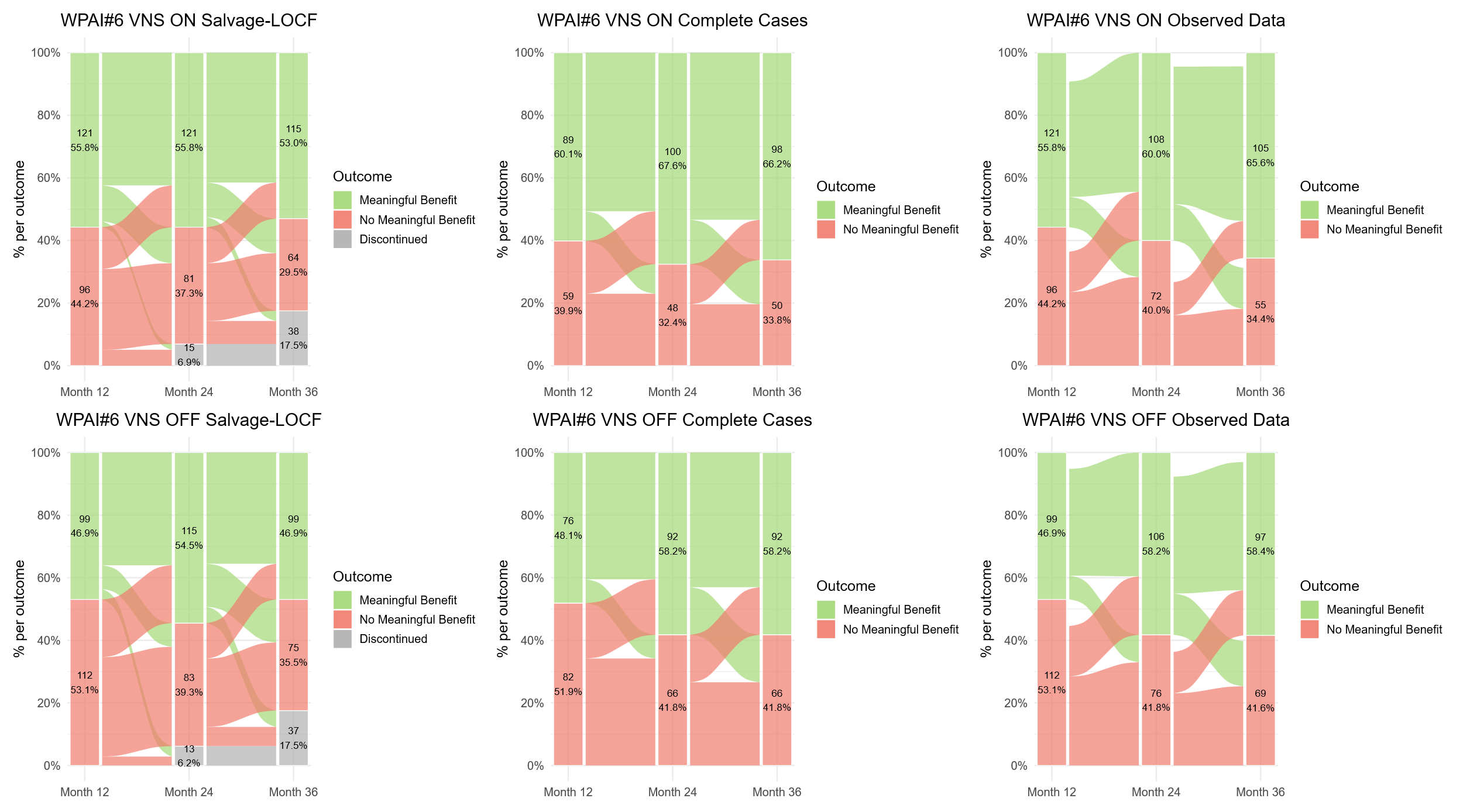 Delayed-Active Group: WPAI Item 6 |
| E. 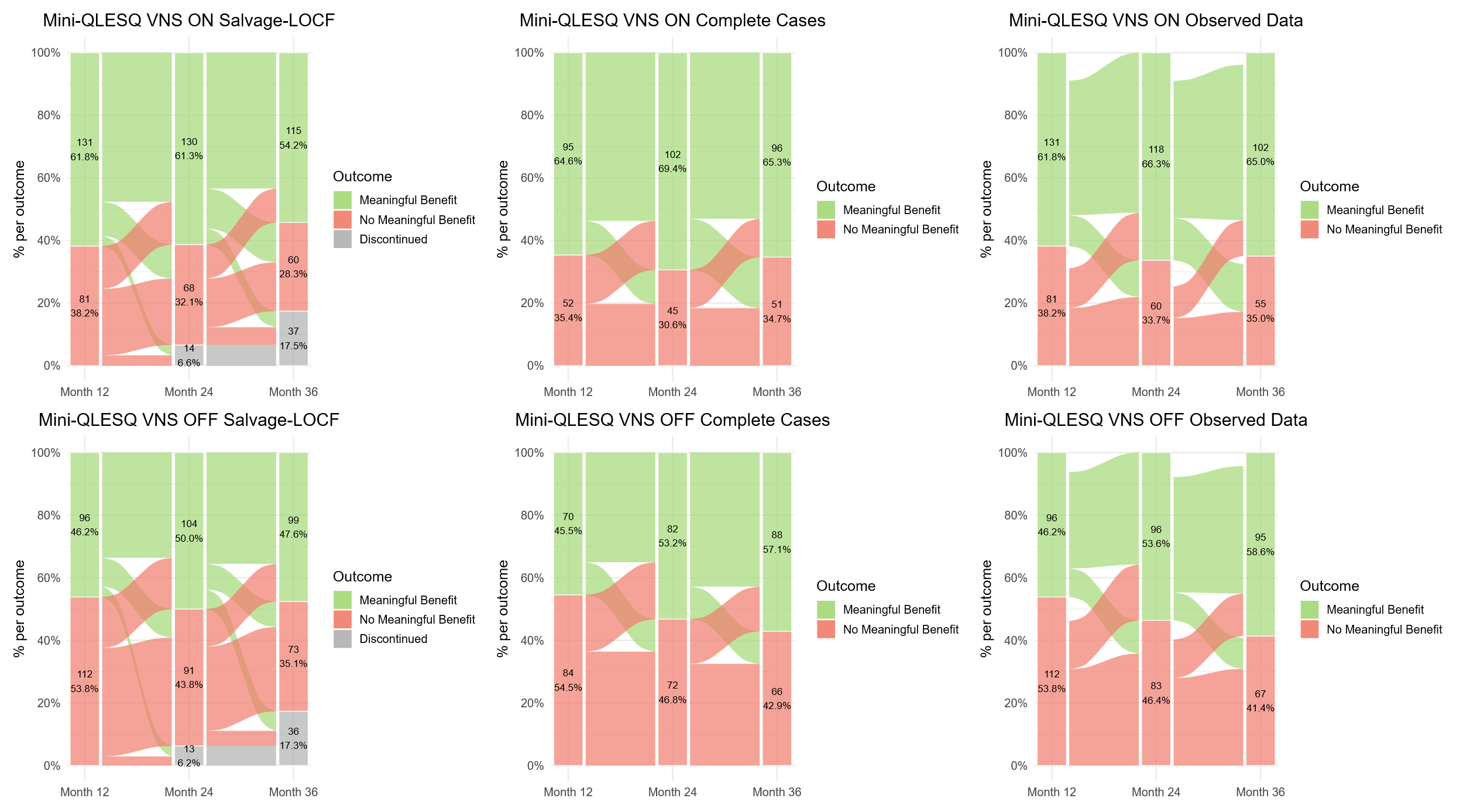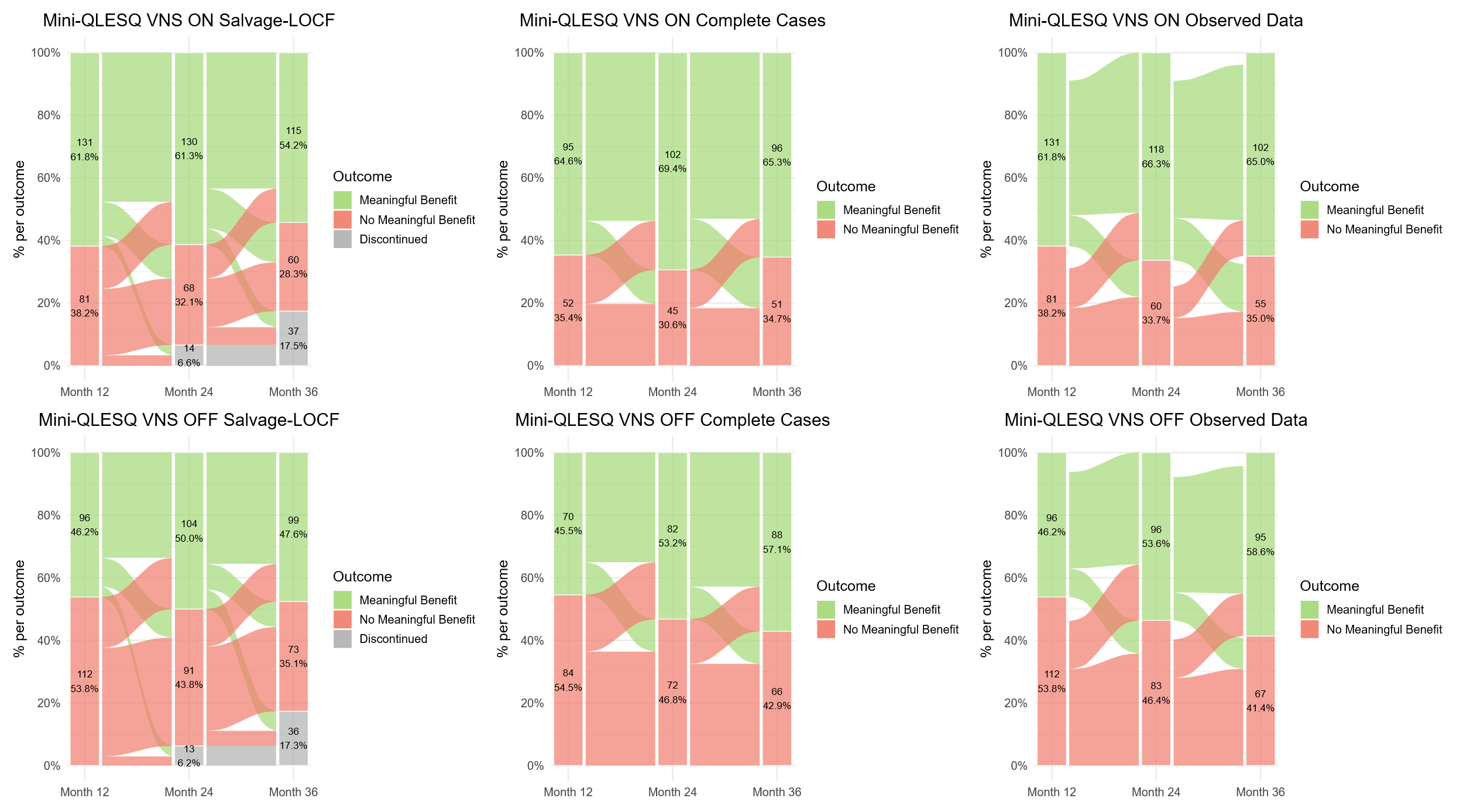Early-Active Group: Mini-Q-LES-Q | F. 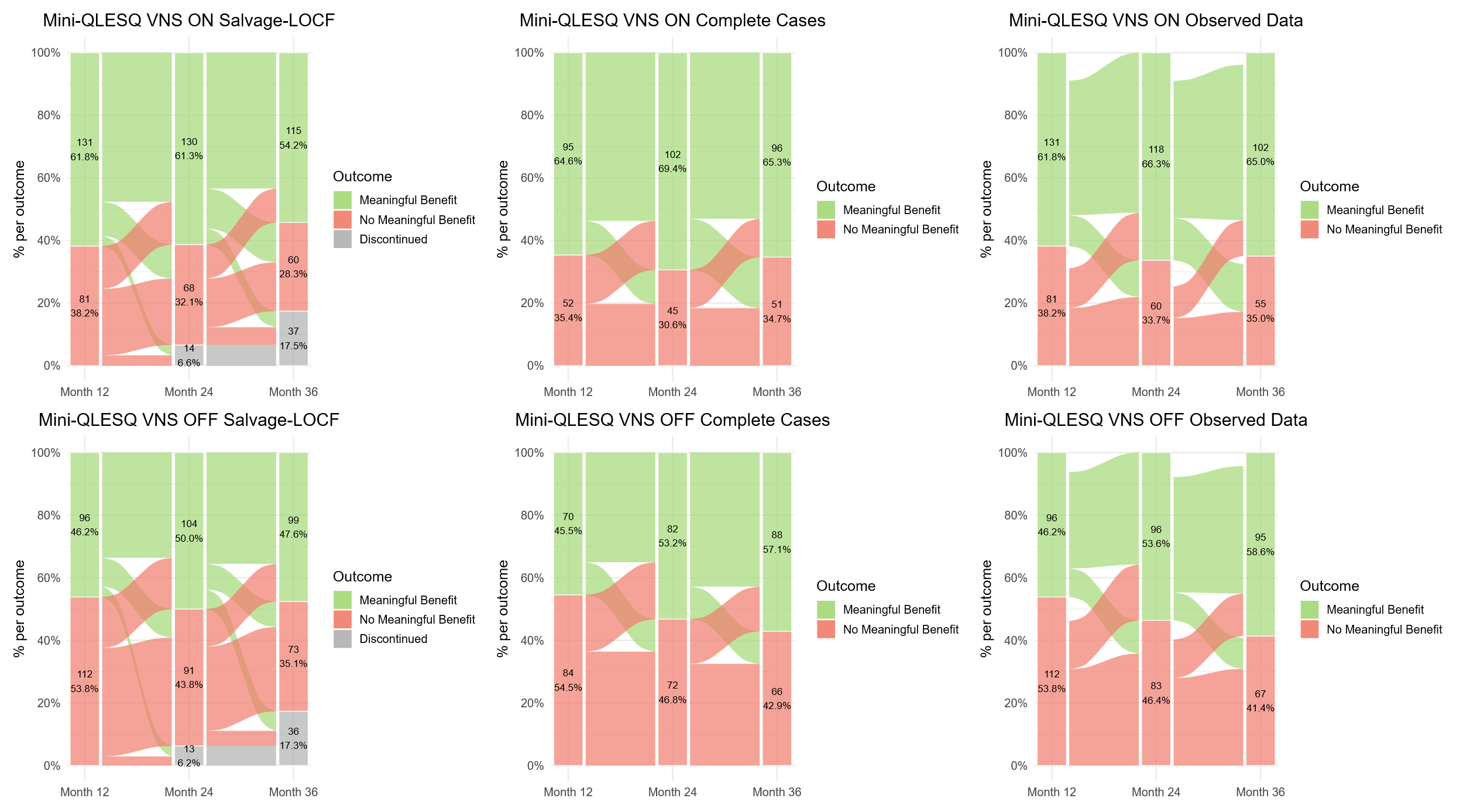Delayed-Active Group: Mini-Q-LES-Q |

Data presented are samples with no missing values at the three time points.

Abbreviations: Mini-Q-LES-Q, 7-item subset of the Quality of Life Enjoyment and Satisfaction Questionnaire; QIDS-SR, Quick Inventory of Depressive Symptomatology–Self-Report; SB, substantial benefit; WPAI, Work Productivity and Activity Impairment Questionnaire.

**Figure S3**. Alluvial diagrams for change in **A., B.** MADRS; **C., D.** QIDS-C; **E., F.** QIDS-SR; and **G., H.** CGI-I outcome categories in the Early-Active and Delayed-Active groups from Month 12 to Month 36 using LOCF. Data are based on exclusive outcome definitions.

| A. 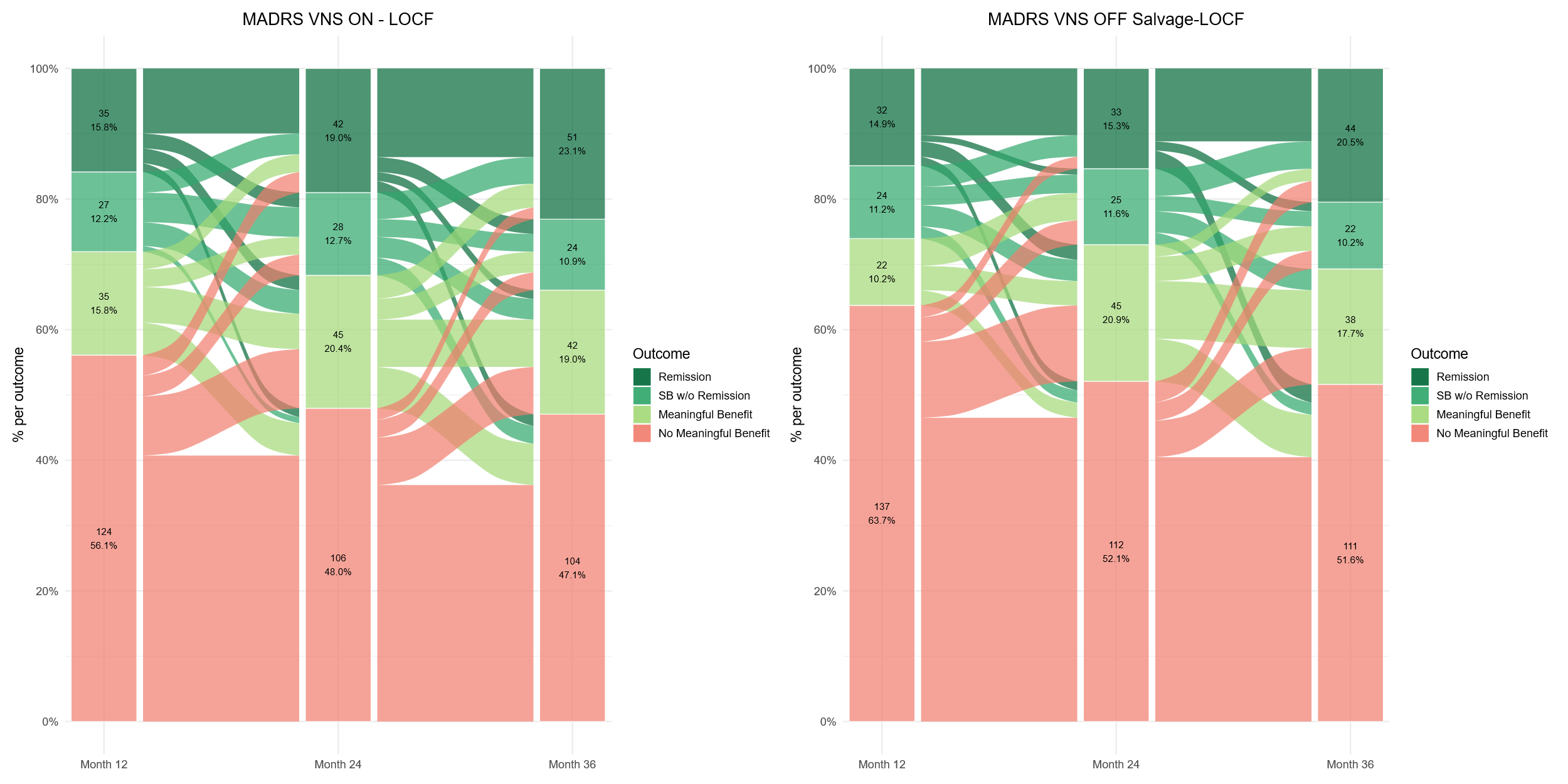 Early-Active Group: MADRS  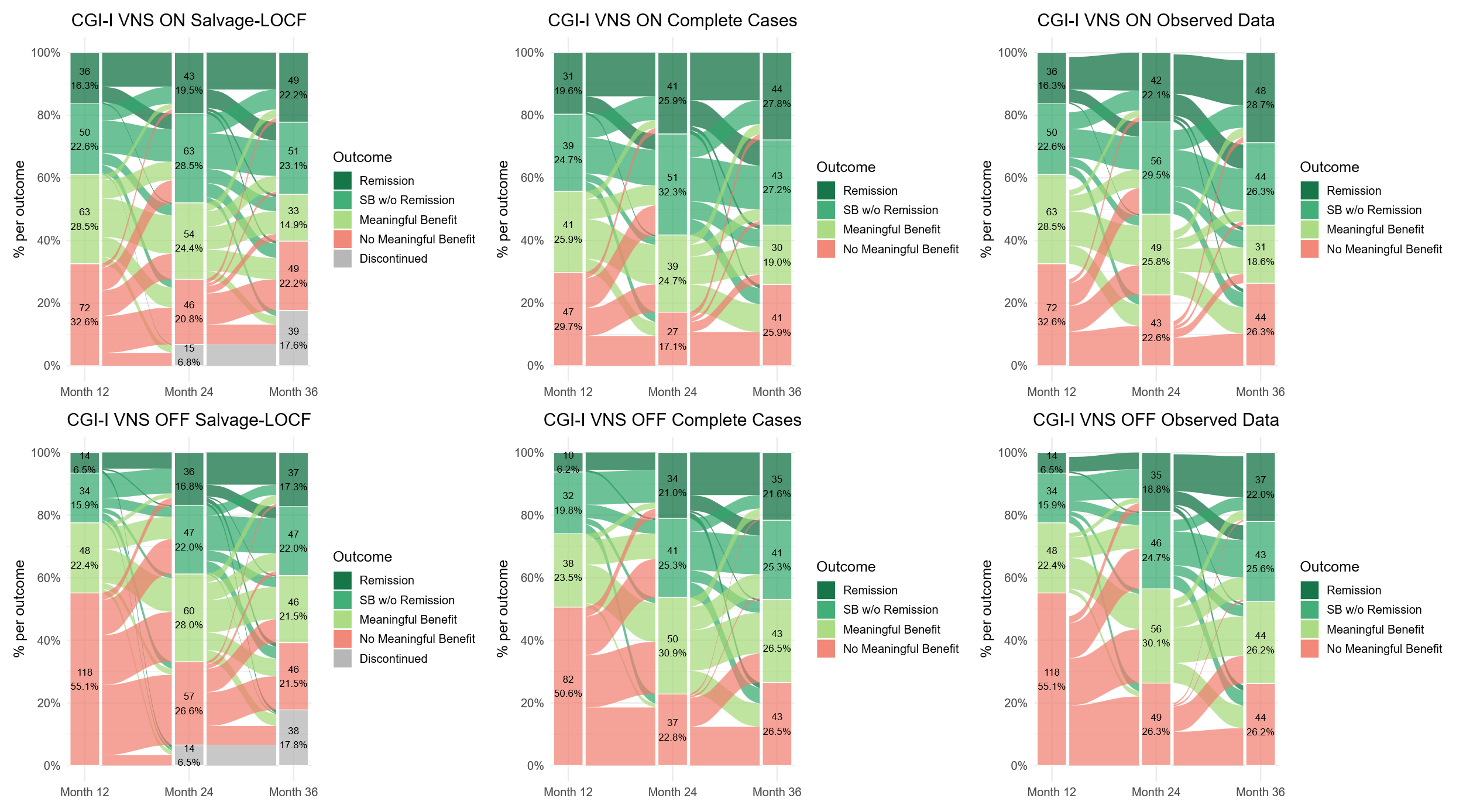 Remission  Response w/o remission  Partial response w/o response  No meaningful benefit | B. 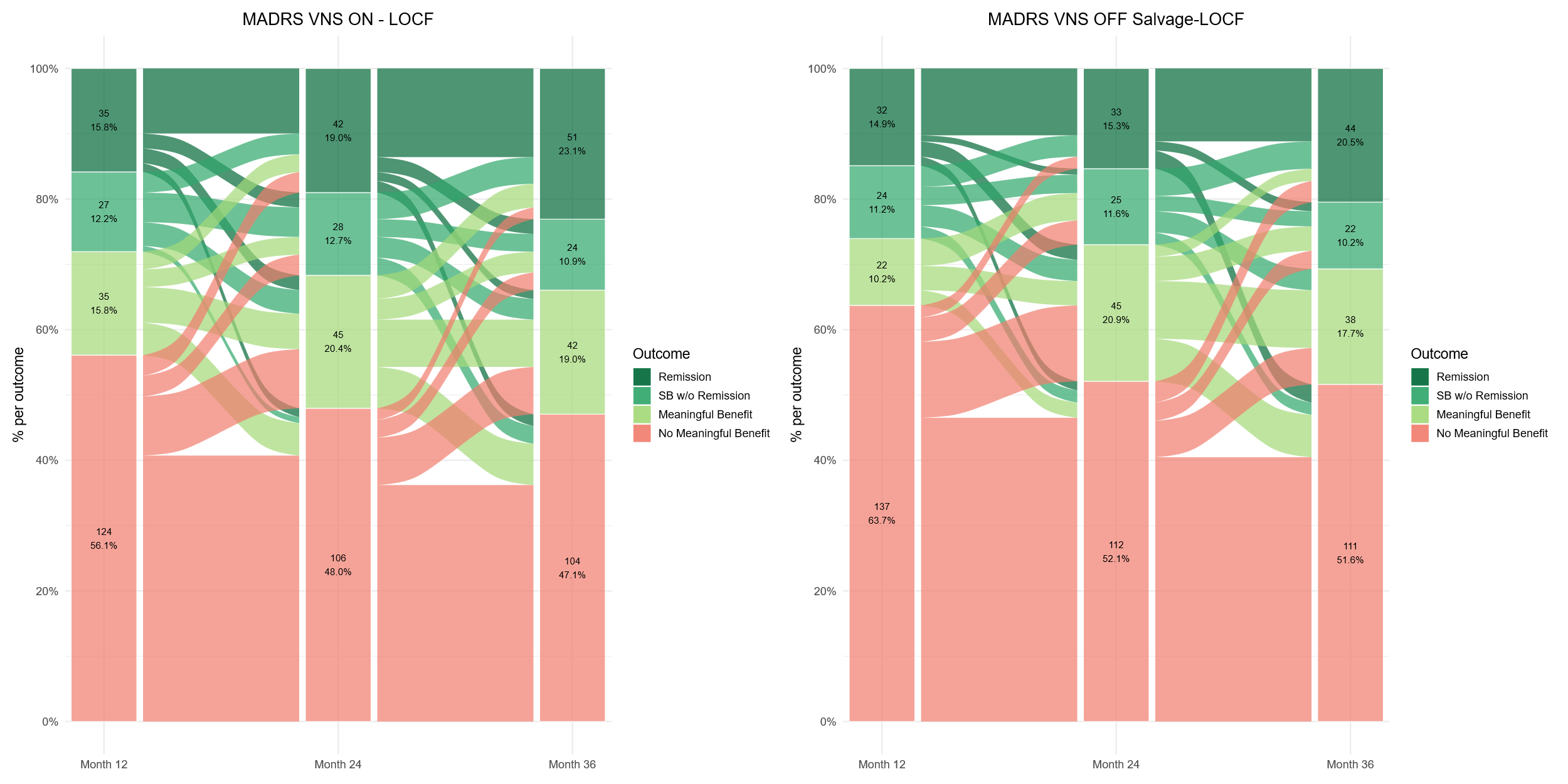 Delayed-Active Group: MADRS |
| --- | --- |
| C. 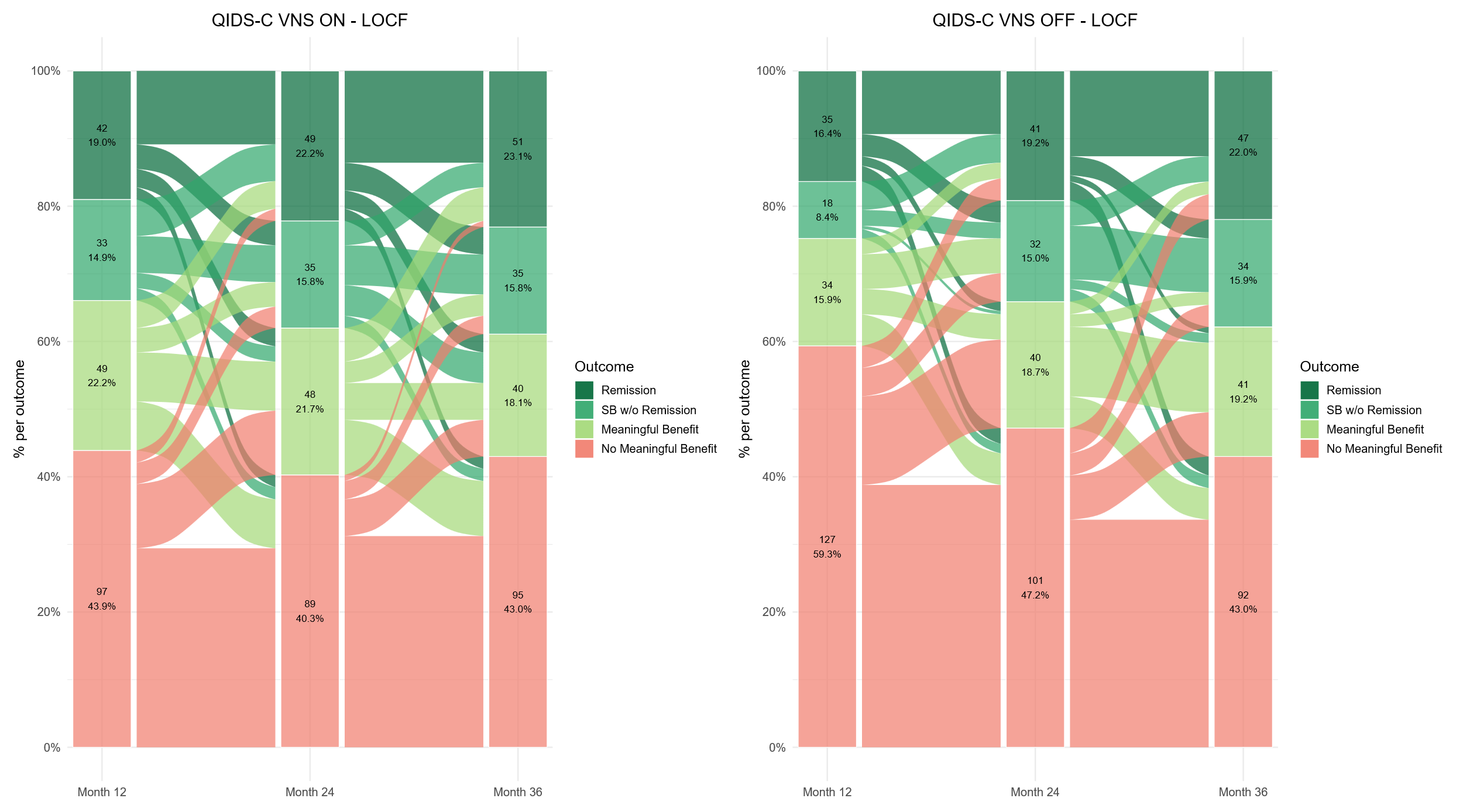 Early-Active Group: QIDS-C | D. 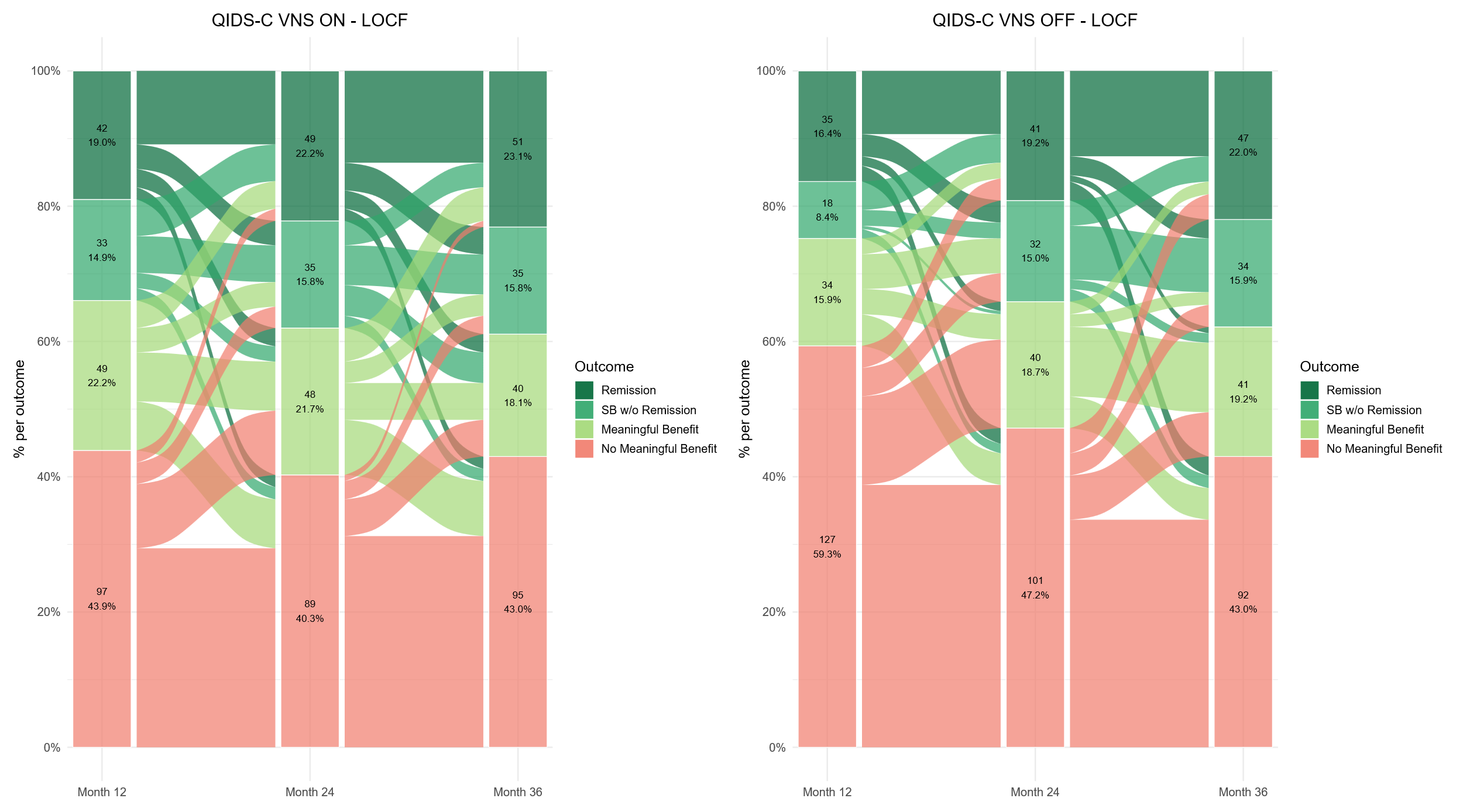 Delayed-Active Group: QIDS-C |
| E. 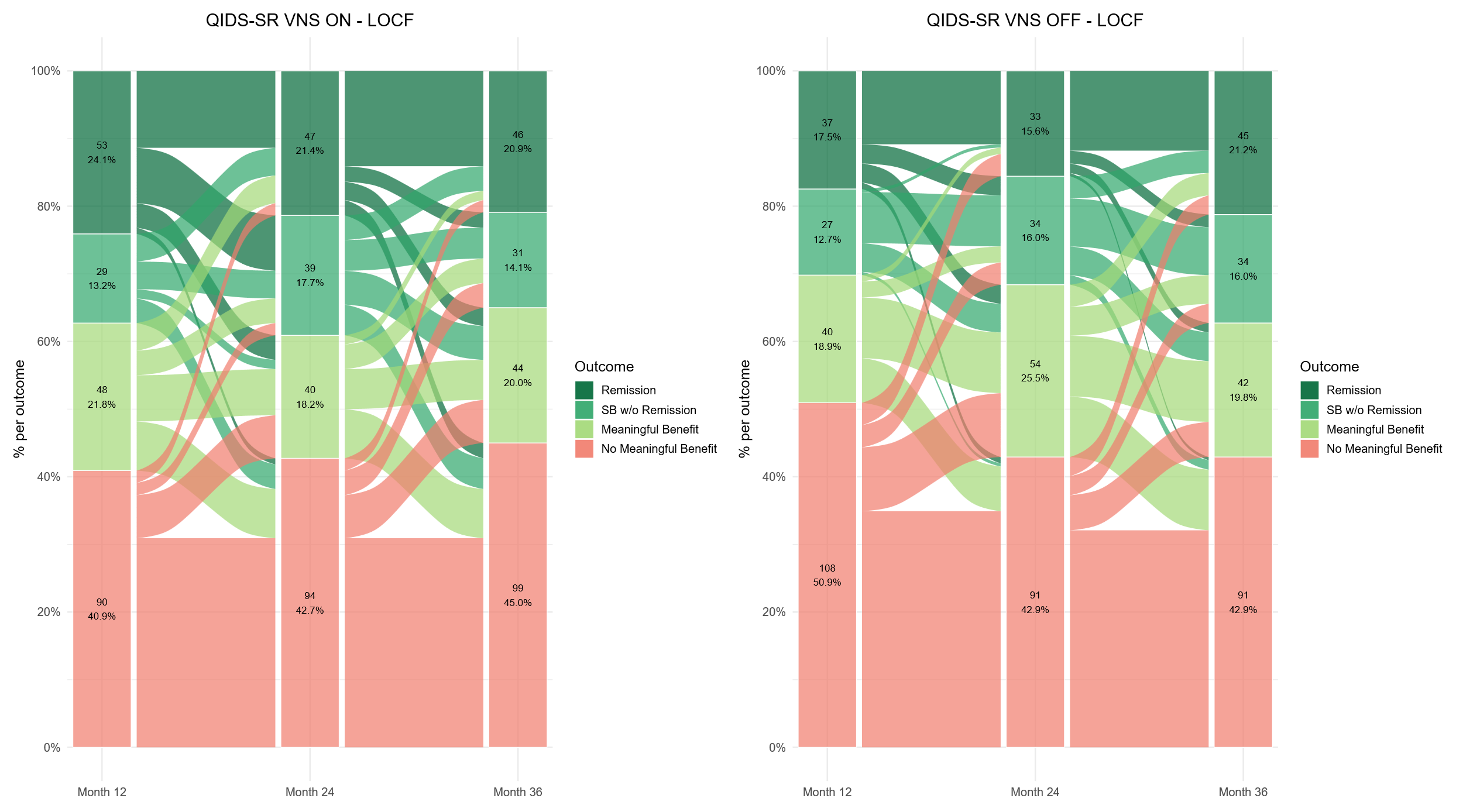Early-Active Group: QIDS-SR  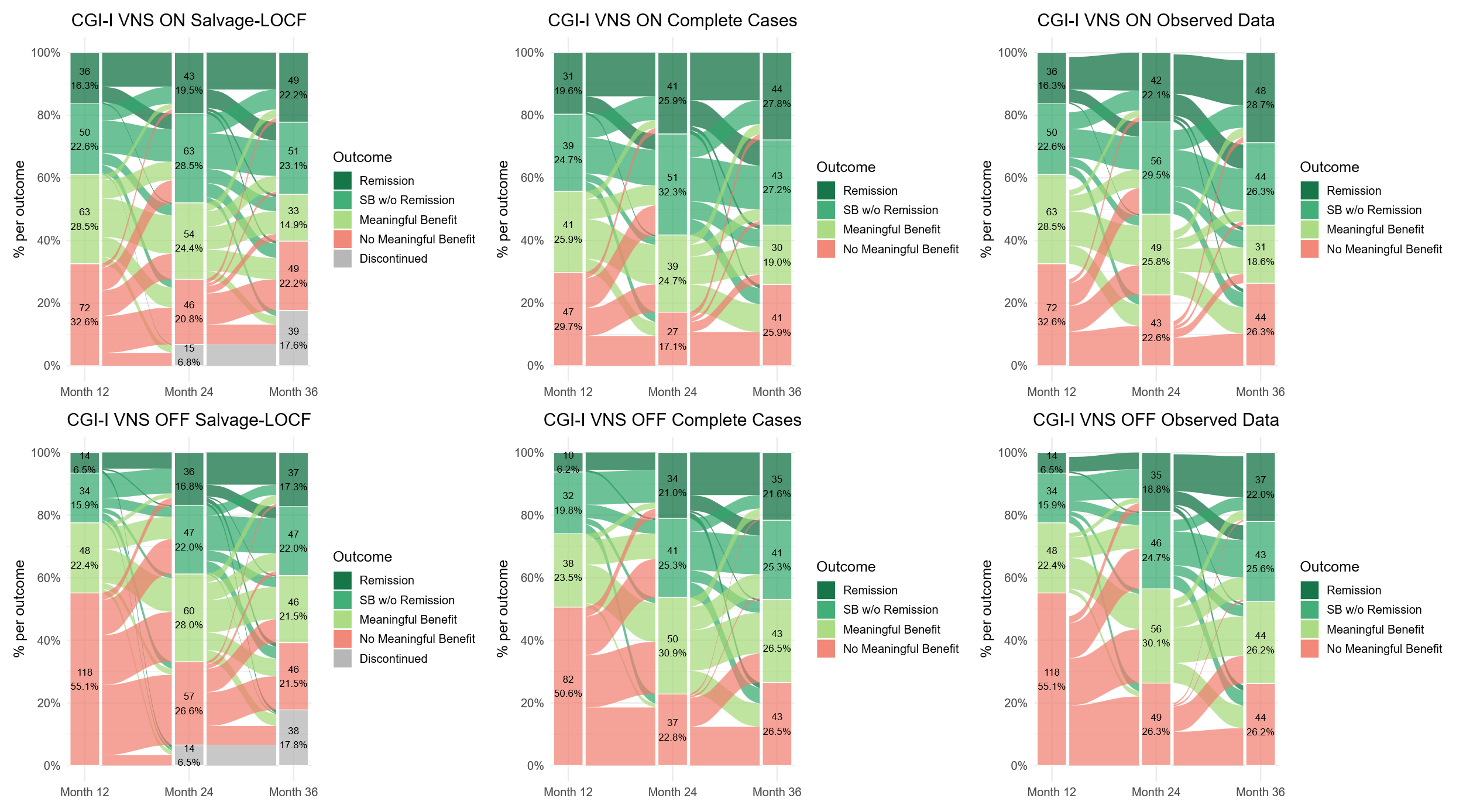 Remission  Response w/o remission  Partial response w/o response  No meaningful benefit | F. 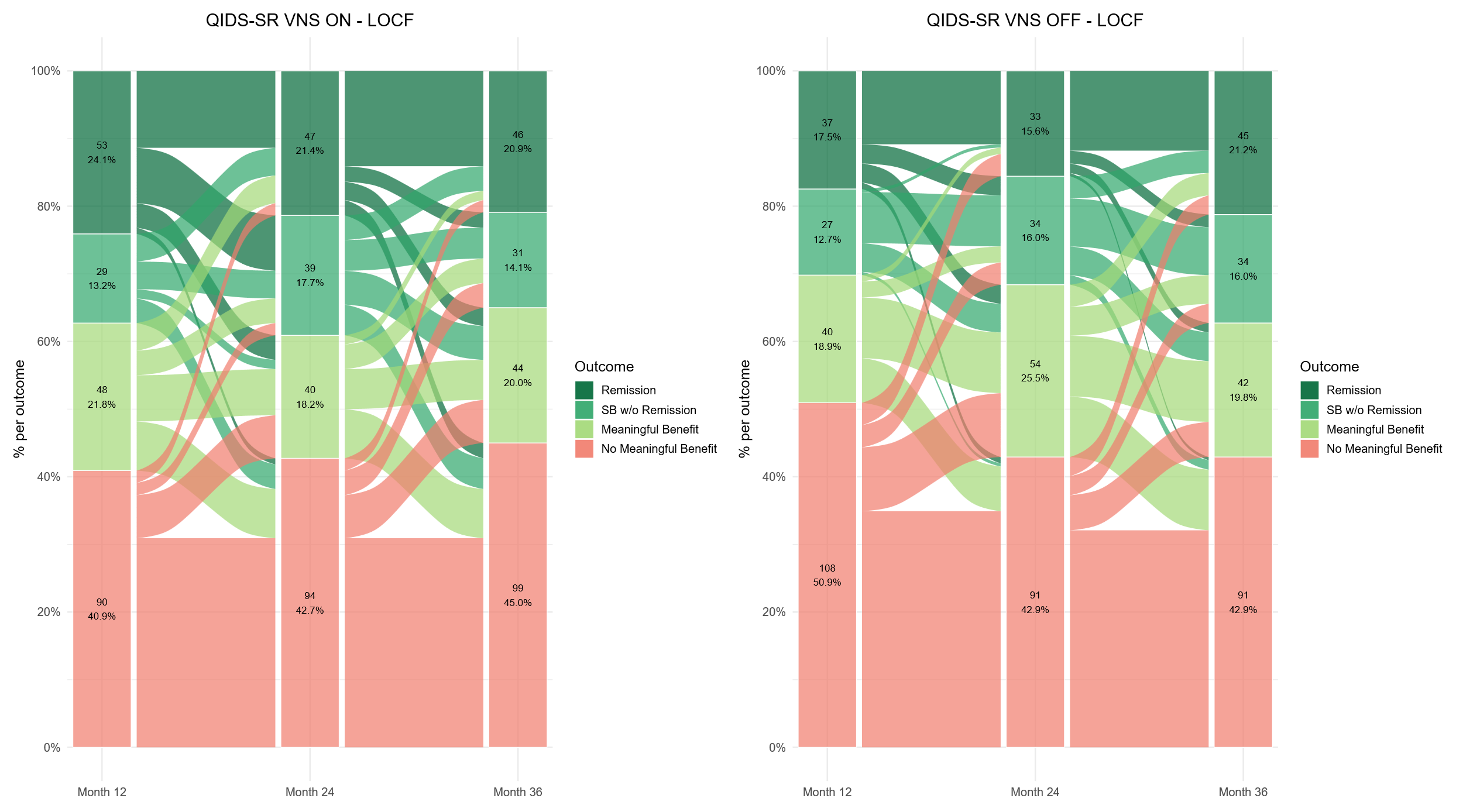Delayed-Active Group: QIDS-SR |
| G. 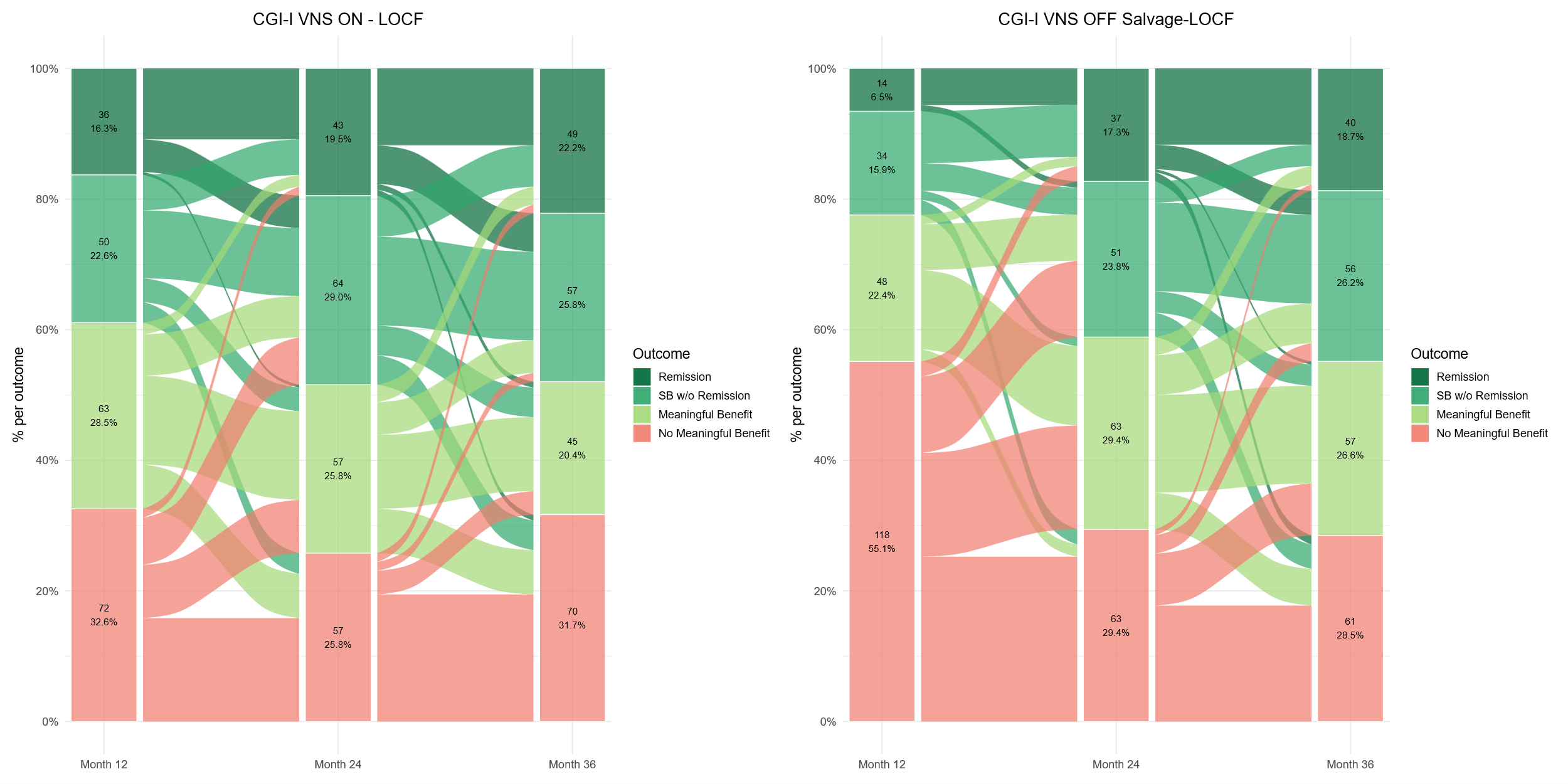Early-Active Group: CGI-I  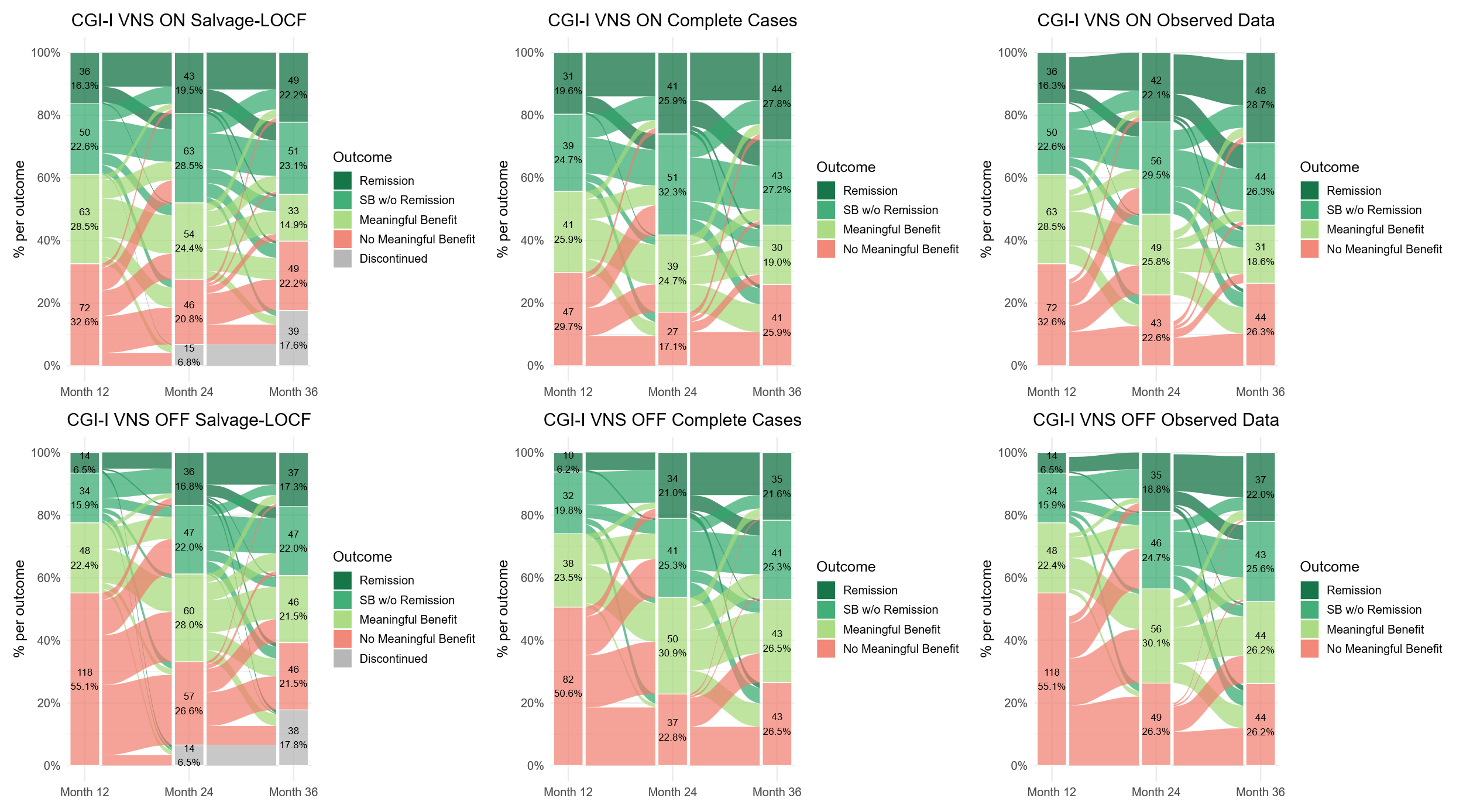 Remission  Response w/o remission  Partial response w/o response  No meaningful benefit | H. 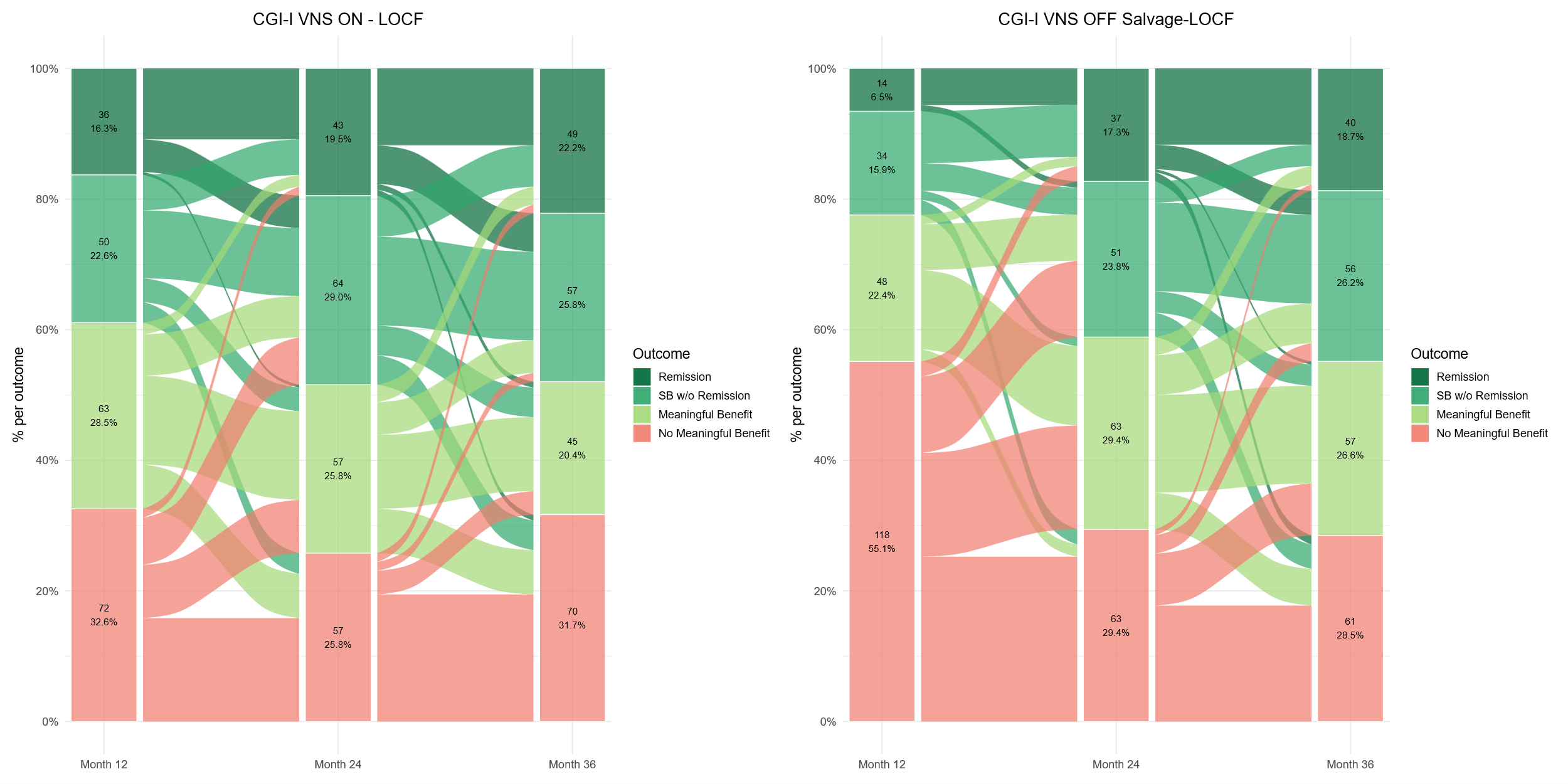Delayed-Active Group: CGI-I |

Data presented use LOCF.

Abbreviations: CGI-I, Clinical Global Impression–Improvement; LOCF, last observation carried forward; MADRS, Montgomery-Åsberg Depression Rating Scale; QIDS-C, Quick Inventory of Depressive Symptomatology–Clinician; QIDS-SR, Quick Inventory of Depressive Symptomatology–Self-Report; SB, substantial benefit.

**Figure S4**. Alluvial diagrams for change in **A., B.** WPAI item 6 and **C., D.** Mini-Q-LES-Q outcome categories in the Early-Active and Delayed-Active groups from Month 12 to Month 36 using LOCF.

| A. 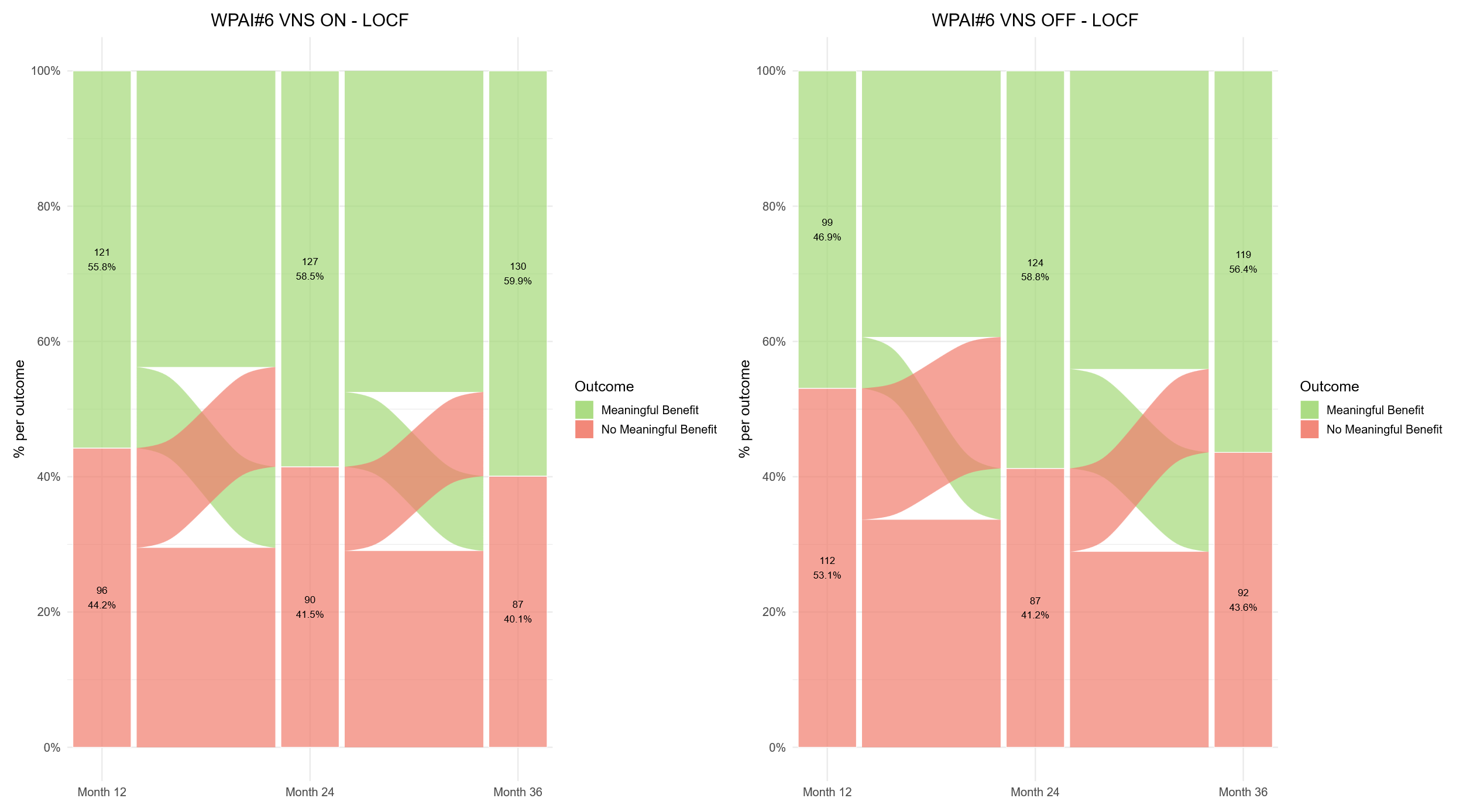Early-Active Group: WPAI Item 6 | B. 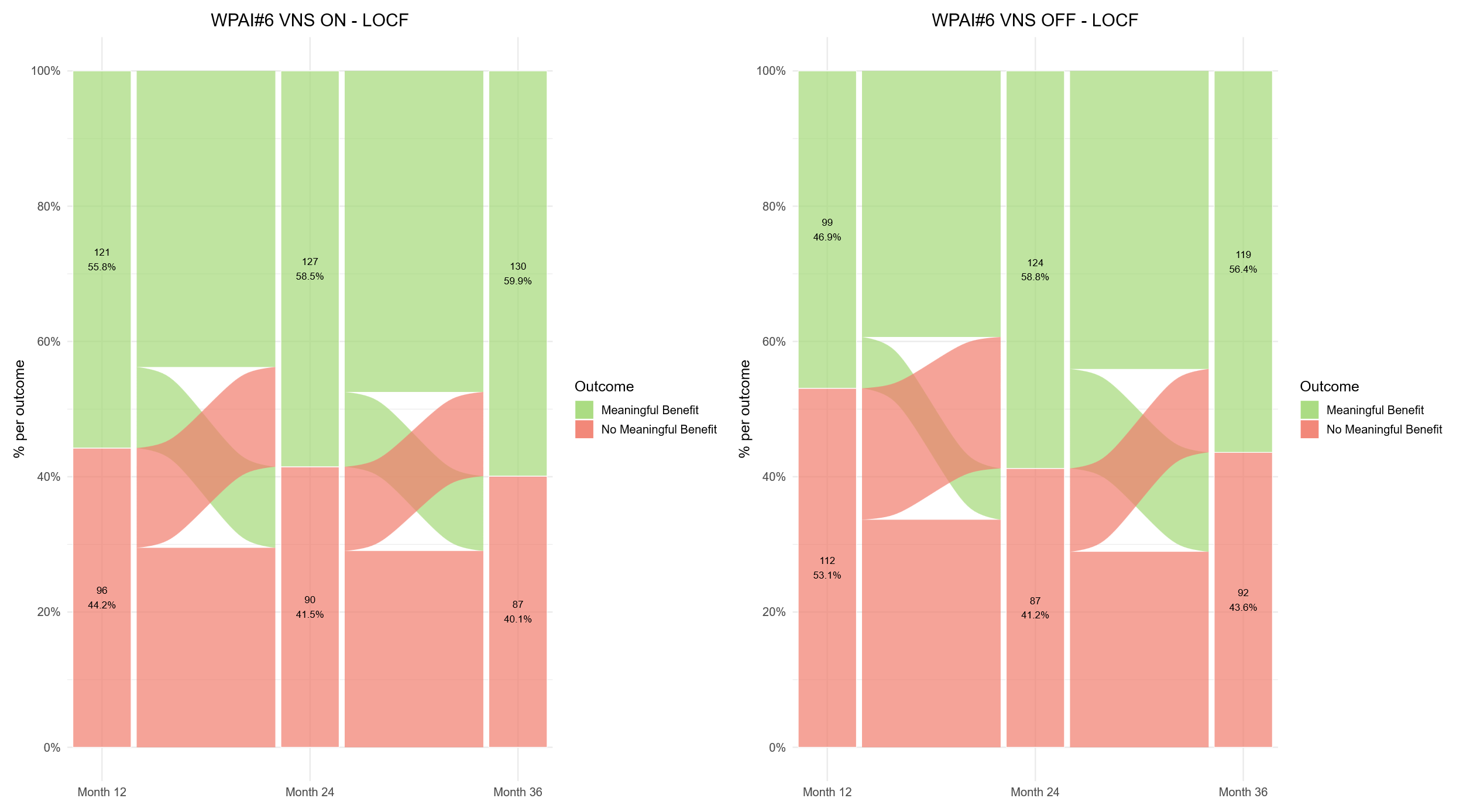 Delayed-Active Group: WPAI Item 6 |
| --- | --- |
| C. 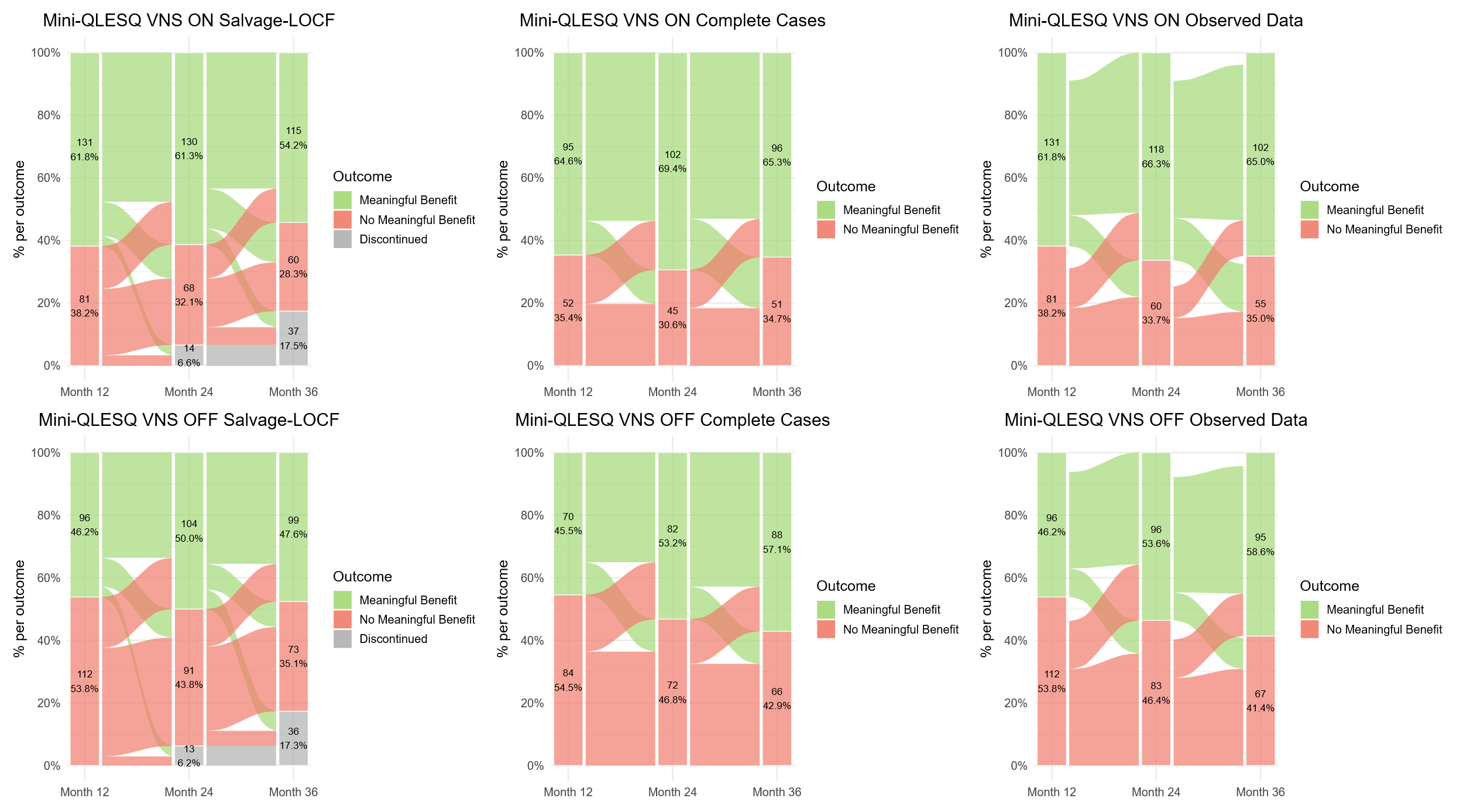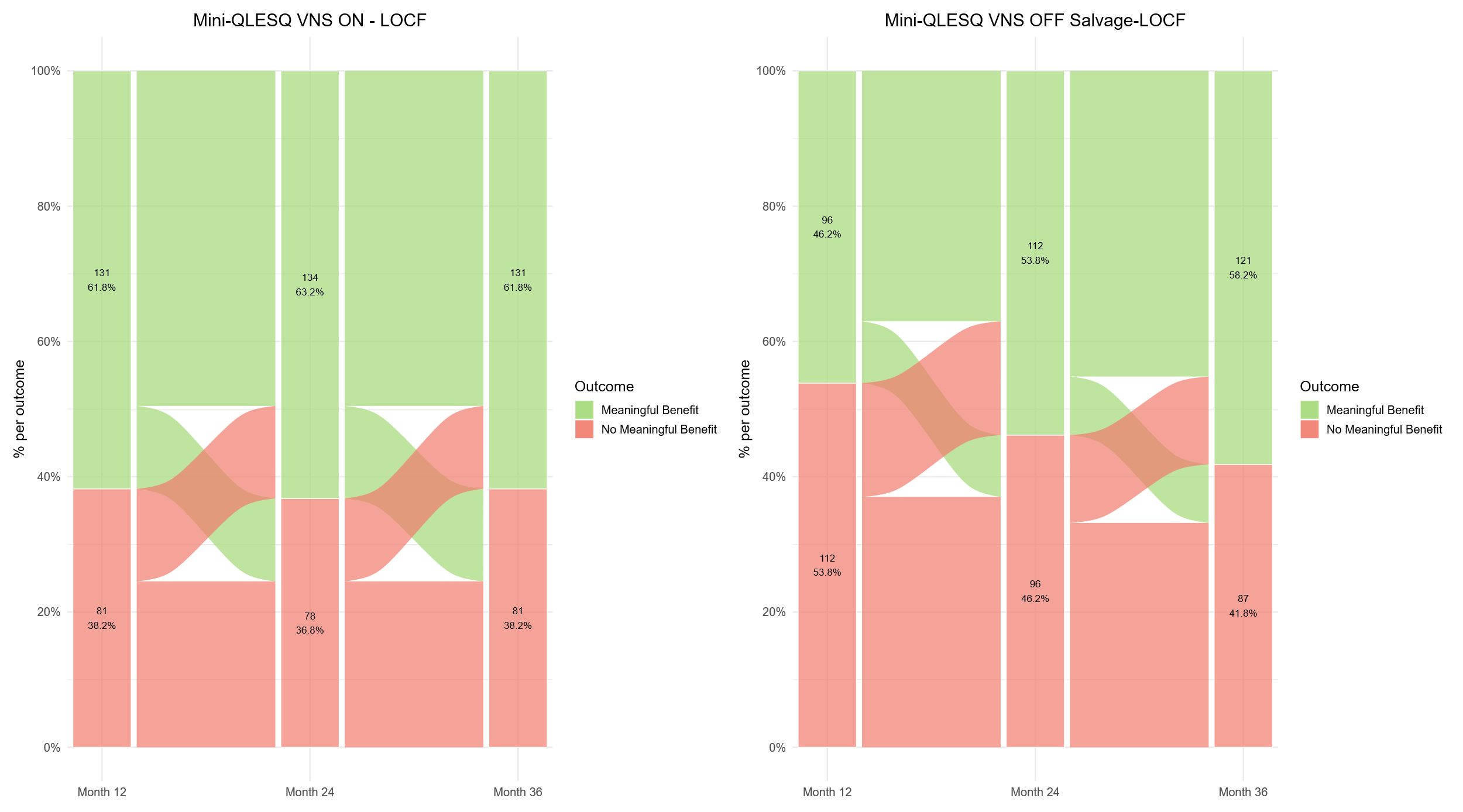Early-Active Group: Mini-Q-LES-Q | D. 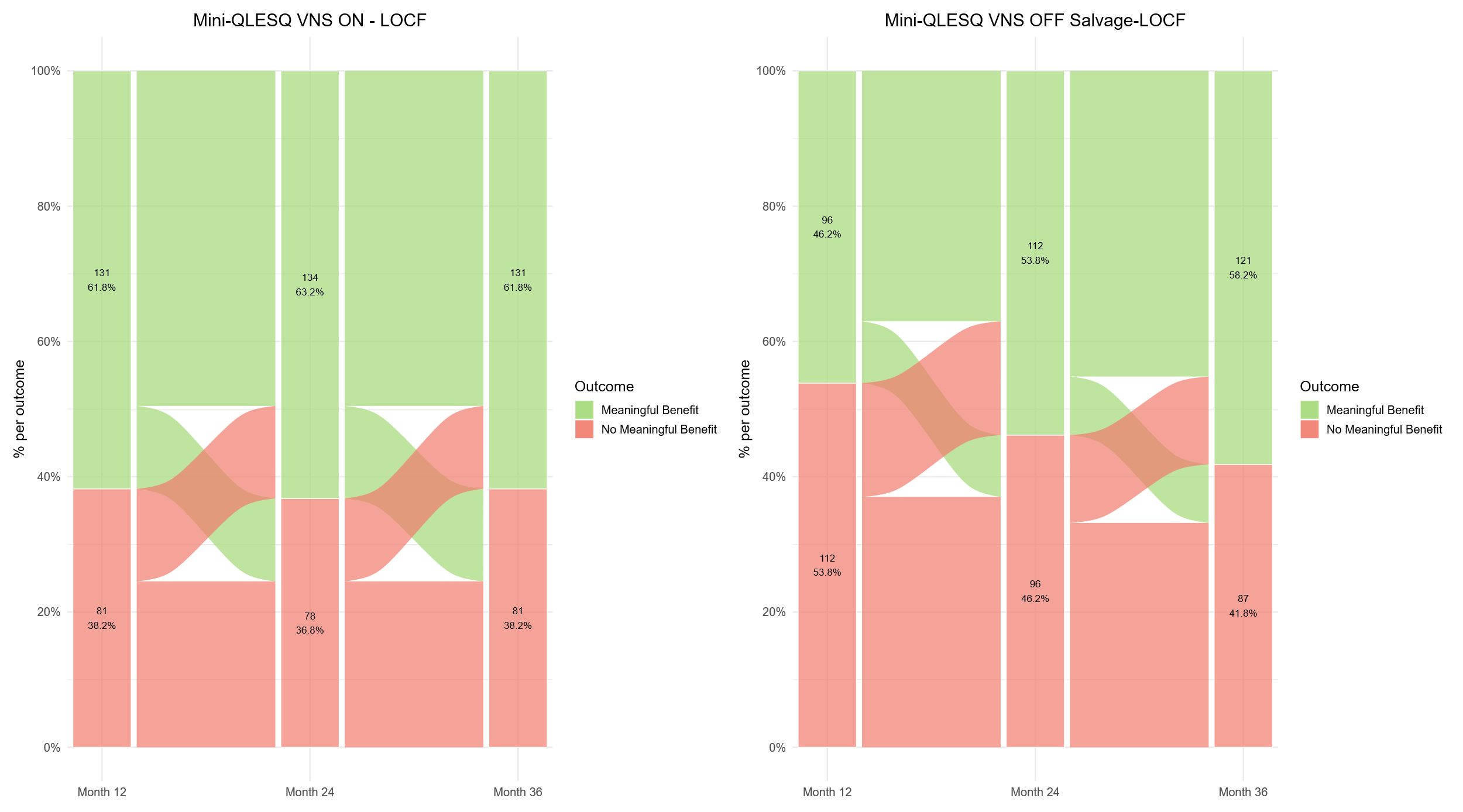Delayed-Active Group: Mini-Q-LES-Q |

Data presented use LOCF.

Abbreviations: LOCF, last observation carried forward; Mini-Q-LES-Q, 7-item subset of the Quality of Life Enjoyment and Satisfaction Questionnaire; WPAI, Work Productivity and Activity Impairment Questionnaire.
